# Poor prognosis of stage I lung adenocarcinoma patients determined by elevated expression over pre/minimally invasive status of COL11A1 and THBS2 in the focal adhesion pathway

**DOI:** 10.1101/2021.12.16.21267913

**Authors:** Jun Shang, Yue Zhao, He Jiang, Jingcheng Yang, Naixin Zhang, Luyao Ren, Qingwang Chen, Ying Yu, Leming Shi, Haiquan Chen, Yuanting Zheng

**Affiliations:** State Key Laboratory of Genetic Engineering, Human Phenome Institute, School of Life Sciences and Shanghai Cancer Center, Fudan University, Shanghai, China; Department of Thoracic Surgery, Shanghai Cancer Center, Fudan University Shanghai, China; Institute of Thoracic Oncology, Fudan University, Shanghai, China; Department of Oncology, Shanghai Medical College, Fudan University, Shanghai, China

**Keywords:** lung adenocarcinoma, pre/minimally invasive, molecular subtypes, prognosis, COL11A1, THBS2, overfitting-resistant, unsupervised clustering

## Abstract

Around 20% of stage I lung adenocarcinoma (LUAD) patients die within five years after surgery, and efforts for developing gene-expression based models for risk-tailored post-surgery treatment are largely unsatisfactory due to overfitting-related lack of validation and extrapolation. Because patients with adenocarcinomas in situ (AIS) and minimally invasive (MIA) LUAD are completely curable by surgical resection, we hypothesize that poor-prognosis stage I patients may exhibit key molecular characteristics deviating from AIS/MIA. We first found focal adhesion (FA) as the only pathway significantly perturbed at both genomic and transcriptomic levels by comparing 98 AIS/MIA and 99 invasive LUAD patients. Then, we identified two FA pathway genes (COL11A1 and THBS2) strongly upregulated from AIS/MIA to stage I while expressed steadily from normal to AIS/MIA. Furthermore, unsupervised clustering separated stage I patients into two molecularly and prognostically distinct subtypes (S1 and S2) based solely on the expression levels of COL11A1 and THBS2 (FA2). Subtype S1 looked like AIS/MIA, whereas S2 exhibited more somatic alterations, elevated expression of COL11A1 and THBS2, and more activated cancer-associated fibroblast (CAF). The prognostic performance of the knowledge-driven and overfitting-resistant FA2 model was validated with 12 external data sets and may help reliably identify high-risk stage I patients for more intensive post-surgery treatment.

## INTRODUCTION

Lung adenocarcinoma (LUAD) is the most common histological subtype of lung cancer with greatly varied five-year survival rate^1,2^. Adenocarcinoma in situ (AIS) and minimally invasive adenocarcinoma (MIA), defined as pre/minimally invasive stage lesions with no or less than 5 mm of invasion, have excellent five-year survival rate of virtually 100%^2-4^. However, the five-year survival rate of invasive LUAD, even in early pathological stage I, drops to about 80%^1^. Meanwhile, there is controversy over whether patients with stage I LUAD benefit from adjuvant therapy^5,6^, causing uncertainties in the clinical treatment of these patients.

There is an urgent need to accurately classify stage I LUAD patients into good-prognosis subgroup like AIS/MIA who can be cured by surgical resection alone and poor-prognosis subgroup with high risk of recurrence or death who may benefit from more aggressive post-surgery treatment such as adjuvant therapy. Over the past 15 years, many predictive models based on gene-expression data from microarray and high-throughput sequencing technologies have been developed for risk stratification of LUAD patients^7^. However, there is still a lack of simple and robust predictive models for molecular subtyping of stage I LUAD suitable for clinical applications^8^. So far, the Oncocyte DetermaRx test (https://oncocyte.com) based on the expression of 14 genes appears to be the only one used in clinic^9^.

The small number of genes (features) and the overfitting-resistant process by which how such genes are selected are two critically important characteristics for the robustness of a predictive model^10^. For most models of molecular classification of stage I LUAD^11,12^, genes are generally selected based on direct comparison of the expression levels between patients with good and bad prognosis. Over-fitting caused by the complex modeling process including the selection of many genes to explicitly fit the prognosis endpoint of the training set, accompanied by inadequate cross-validation and external validation with truly independent data sets, is an important reason why many published models have not been adopted for clinical applications^8,10^. In addition to risk stratification, understanding the molecular characteristics and tumor microenvironment of subtypes of stage I LUAD may help lay the foundation for precision patient treatment.

The molecular alterations from AIS/MIA to invasive LUAD can provide useful insight beyond the degree of pathological invasion for better understanding the disease but have not been adequately studied. Thus, in a previous study^13^, we comprehensively analyzed the genomic and immune profiling of AIS/MIA and invasive LUAD, and identified potential driver mutation events such as TP53 mutation, arm-level copy number variation (CNV), and HLA loss of heterozygosity. However, details about the differentially expressed or mutated genes (DEGs or DMGs) between AIS/MIA and invasive LUAD remain to be fully explored. We hypothesize that such DEGs and DMGs, when combined, may help identify key genes associated with the early progression to stage I LUAD from AIS/MIA, and such key genes may then be used to further stratify stage I LUAD patients into subtypes with divergent molecular characteristics and prognosis, in a completely knowledge-driven, unsupervised and robust manner. Partition Around Medoids (PAM) ^14,15^, which is well-known for its robust performance in identifying the true underlying number of clusters within a data set by resisting noise and isolated data points, may serve this purpose.

In this study, we successfully revealed two molecularly and prognostically distinct subtypes of stage I LUAD by applying key molecular alterations from AIS/MIA to invasive LUAD progression. First, the focal adhesion (FA) pathway and associated DEGs were identified after we thoroughly compared the differences in genomics and transcriptomics between AIS/MIA and invasive LUAD, without any training based on patient prognosis information. Secondly, stage I LUAD patients were further clearly separated into two subtypes (S1 and S2) based on a simple unsupervised partition model using the expression levels of only two genes (COL11A1 and THBS2) in the FA pathway. Thirdly, we comprehensively analyzed the molecular characteristics of S1 and S2. S1 was closer to pre/minimally invasive LUAD in genomics, transcriptomics, and tumor microenvironment; whereas subtype S2 demonstrated elevated expression of COL11A1 and THBS2, higher somatic alterations, and more active in cancer-associated fibroblast (CAF), diverging markedly from pre/minimally invasive LUAD. Finally, 12 published data sets consisting of 1,368 stage I LUAD patients validated our findings that S2 patients had much worse prognosis than S1 patients.

## RESULTS

### Study design and workflow

The study design and workflow is shown in **Figure 1**. Briefly, a total of 197 patients with primary tumor tissues and matched normal tissues were enrolled in this study at Fudan University Shanghai Cancer Center (FUSCC). Among them, 98 were AIS (24) or MIA (74) LUAD, termed pre/minimally invasive, and 99 were stage I (83) or IIIA (16) invasive LUAD, termed invasive, according to the 7^th^ TNM staging (**Figure 1A**). RNA sequencing (RNA-seq) and whole-exome sequencing (WES) were carried out for the 197 (98+99) tumor-normal pairs of samples (**Figure 1B**). First, we identified 264 DEGs and 25 DMGs between pre/minimally invasive and invasive LUAD corresponding to the transcriptomic and genomic alterations, respectively (**Figure 1C**). Secondly, we identified 11 and 35 pathways that were enriched with the DEGs and DMGs, respectively. Strikingly, focal adhesion (FA), involving a total of 199 genes including 15 DEGs and 1 DMGs (with an overlap of COL11A1), was the only pathway that was significantly perturbed both transcriptomically and genomically (**Figure 1C**). Thirdly, two genes (COL11A1 and THBS2) that were most significantly differentially expressed between invasive and AIS/MIA, and that at the same time showed no difference in expression between AIS/MIA and normal, were identified to develop a model (FA2) using an unsupervised consensus clustering method called Partition Around Medoids (PAM), clearly separating stage I invasive LUAD patients into two distinct subtypes (S1 and S2) (**Figure 1D**). Finally, the differences between subtypes S1 and S2 in molecular characteristics including genomics, transcriptomics, proteomics, tumor microenvironment and clinical outcomes were comprehensively evaluated and confirmed with multiple data sets previously reported in the literature (**Figure 1E**).

**Figure 1.**
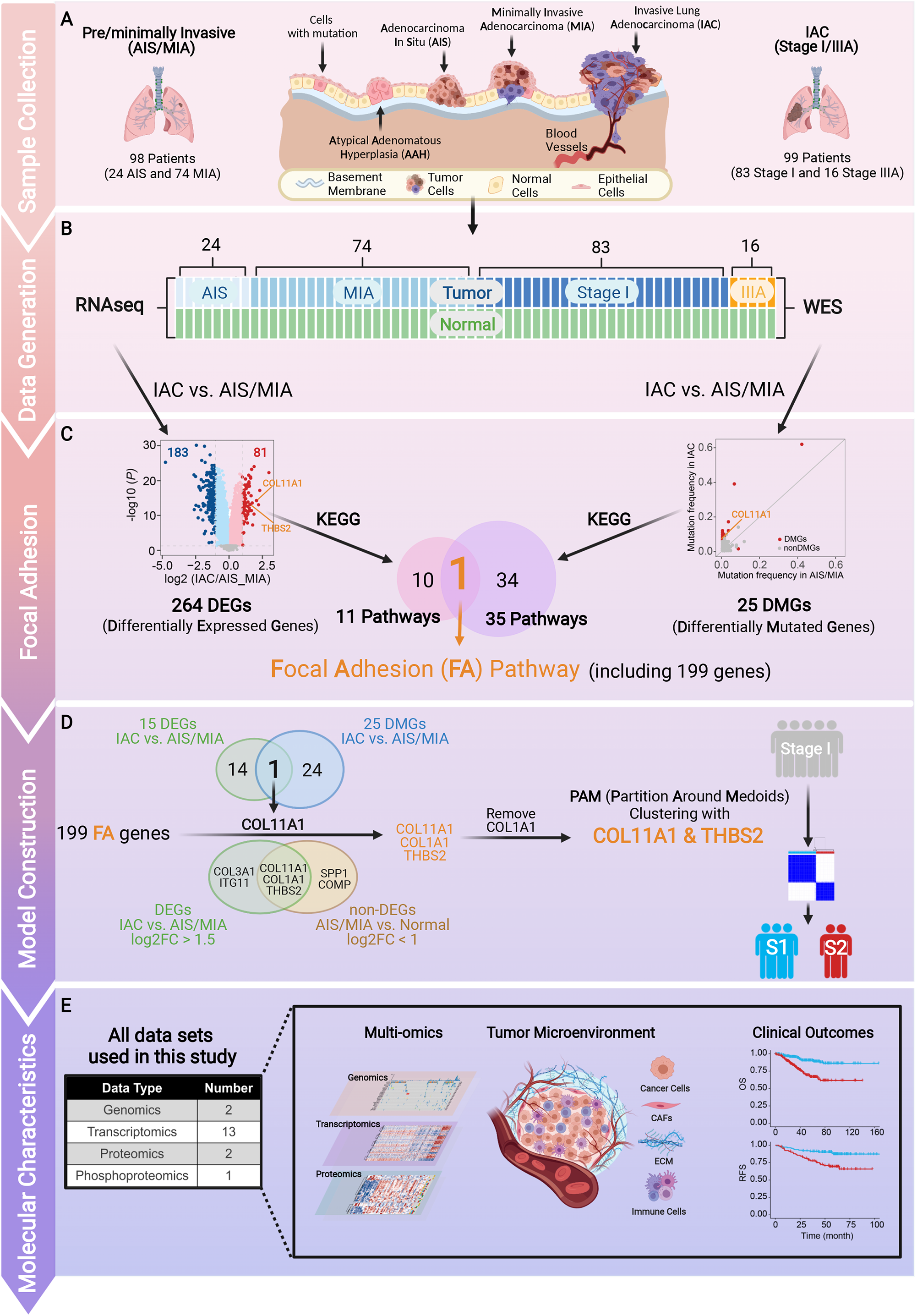
Workflow of identification of stage I LUAD subtypes and associated molecular characteristics. (A) Patients with AIS/MIA and invasive LUAD were enrolled and (B) tissue samples were collected for sequencing with WES and RNA-seq. DMGs and DEGs between AIS/MIA and invasive LUAD were identified. (C) DMGs and DEGs were both enriched in the focal adhesion (FA) pathway by KEGG enrichment analysis. (D) COL11A1 and THBS2 were retained to construct the clustering model, and the PAM consensus clustering using expression of COL11A1 and THBS2 classified stage I LUAD into subtypes S1 and S2. (E) Extensive differences in multi-omics molecular characters, tumor microenvironment (TME), and clinical outcomes between subtypes S1 and S2 were explored in the FUSCC and external data sets.

### Genomic and transcriptomic alterations from pre/minimally invasive to invasive LUAD identified focal adhesion (FA) pathway and elevated expression of COL11A1 and THBS2 as key changes of LUAD progression

AIS and MIA are similar in genomic and transcriptomic characters. The principal component analysis (PCA) suggested that the expression profiles of AIS and MIA were similar to each other and were closer to that of invasive LUAD than to normal lung tissue (**Figure 2A**). Meanwhile, almost no significant DMGs and DEGs between AIS and MIA could be identified (**Figures S1A** and **S1B**). Therefore, we combined AIS and MIA into one group (AIS/MIA) in the following analyses.

**Figure 2.**
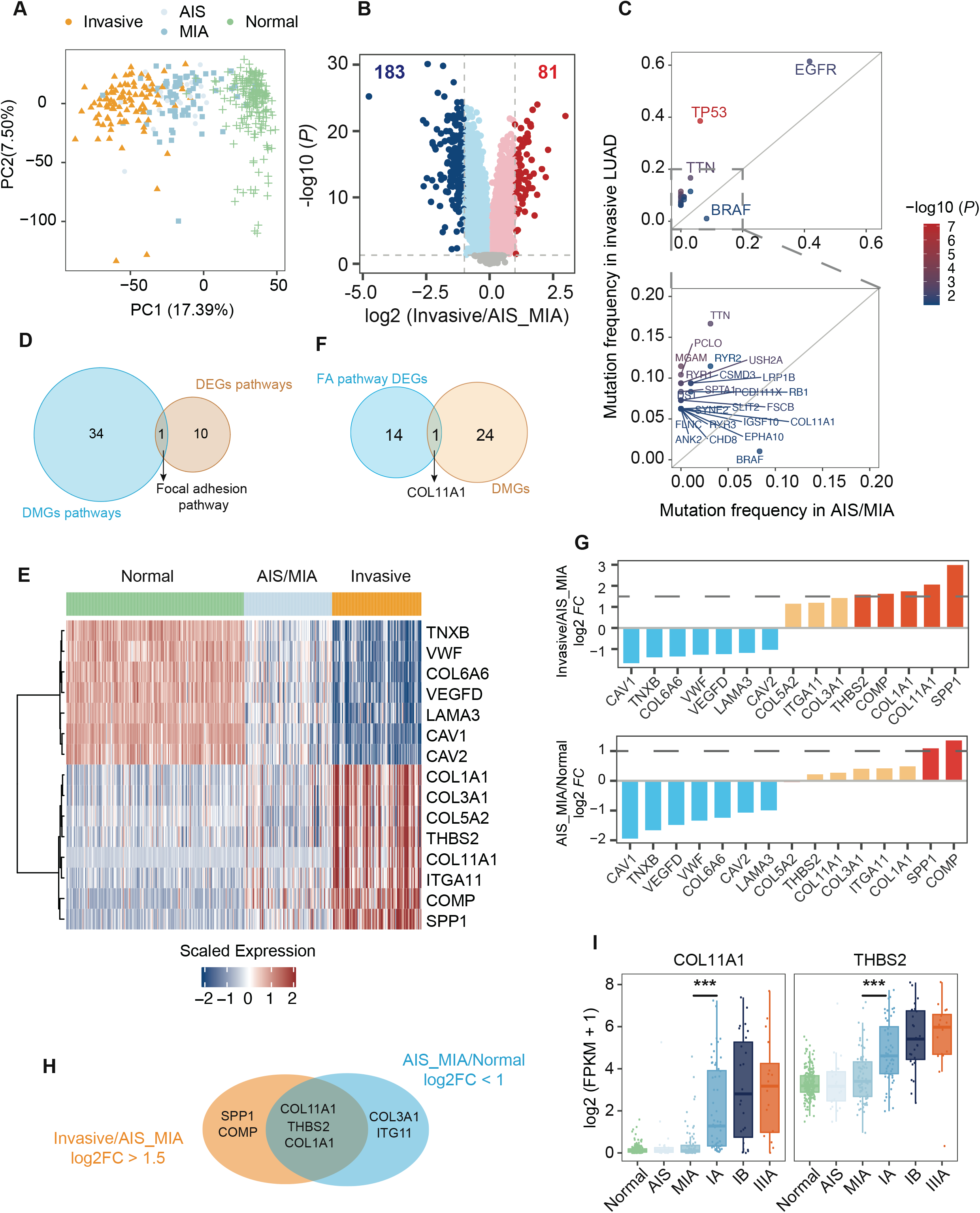
Identification of COL11A1 and THBS2 in the FA pathway as key determinants for invasive LUAD deviating from pre/minimally invasive status. (A) PCA of the expression profiles of 39,476 genes in 197 pairs of LUAD samples including 24 pairs of AIS, 74 pairs of MIA, and 99 pairs of invasive LUAD. (B) Volcano plot shows differential gene expression between invasive LUAD and AIS/MIA. (C) Comparison of gene mutation frequency between AIS/MIA and invasive LUAD. A total of 25 genes show significantly different mutation frequencies between AIS/MIA and invasive LUAD. Color bar shows -log10 (*P*). (D) Venn diagram shows the intersection of pathways enriched by DMGs and DEGs. DMGs and DEGs were both enriched in the FA pathway. (E) The expression of 15 DEGs in the FA pathway between AIS/MIA and invasive LUAD. (F) Venn diagram shows the intersection of 15 DEGs in the FA pathway and 25 DMGs between AIS/MIA and invasive LUAD. (G) The log2FC of the 15 DEGs in the FA pathway: invasive vs AIS/MIA (top) and AIS/MIA vs normal (bottom). (H) Venn diagram shows genes with expression significantly increased from AIS/MIA to invasive LUAD, but no significant difference between AIS/MIA and normal. (I) The expression of COL11A1 and THBS2 from normal to stage IIIA. (*** *P* < 0.001).

We set out to determine important and reliable disease progression-associated pathways by first detecting DEGs and DMGs between AIS/MIA and invasive LUAD. We thus identified 264 DEGs (|log2FC| >=1 and *P* < 0.05) and 25 DMGs (*P* < 0.05) (**Figures 2B, 2C, S1C** and **S1D**). Except for BRAF (AIS/MIA vs invasive, 8% vs 1%), the other 24 DMGs, such as TP53 (AIS/MIA vs invasive, 6% vs 38%), showed much higher mutation frequency in invasive LUAD than in AIS/MIA (**Figures 2C, S1C** and **S1D**). We performed Kyoto Encyclopedia of Genes and Genomes (KEGG) enrichment analysis using the 264 DEGs and 25 DMGs separately. As a result, the focal adhesion (FA) pathway was commonly shared by the 11 DEG-enriched pathways and the 35 DMG-enriched pathways (**Figures 2D, S1E** and **S1F**). It has been reported that the FA complex is a bridge between cells and the extracellular matrix, and plays an important role in cell proliferation, invasion, and migration^16^. We further identified 15 DEGs from the 199 FA pathway genes, which were downloaded from the molecular signature database (MsigDB) (**Figure 2E**). COL11A1, which was the only gene shared by the 15 DEGs and the 25 DMGs in the FA pathway, may play an important role in the progression of AIS/MIA to invasive LUAD (**Figure 2F**).

We hypothesize that genes whose expression is significantly increased only from AIS/MIA to invasive (corresponding to good and bad prognosis, respectively), but not from normal to AIS/MIA (both with good prognosis), may play a more prominent role in disease progression and prognosis. By setting a more stringent log2FC > 1.5 cutoff, we retained five genes (SPP1, COL11A1, COL1A1, COMP, and THBS2) with significantly increased expression level from AIS/MIA to invasive. However, two (SPP1 and COMP) of them had already shown significantly higher expression level in AIS/MIA than in normal, and therefore were removed from further consideration (**Figure 2G**). This process led to three remaining genes, namely COL11A1, THBS2, and COL1A1 (**Figure 2H**). Considering that COL1A1 and COL11A1 are from the same gene family with similar functions, we only used COL11A1 and THBS2 for subsequent molecular subtyping analysis of stage I LUAD. Obviously, we can see that the expression levels of COL11A1 and THBS2 both increased significantly from normal/AIS/MIA to stage IA (**Figure 2I**), whereas no appreciable change in expression was observed from normal to AIS to MIA.

### Unsupervised consensus clustering classified stage I LUAD into AIS/MIA-like subtype S1 and AIS/MIA-diverging subtype S2 based solely on the expression of COL11A1 and THBS2

We assumed that stage I LUAD patients may be further divided into multiple molecular subtypes including those similar or dissimilar to AIS/MIA in different degrees based on the expression of the two FA genes (FA2). Thus, we used the unsupervised Partition Around Medoids (PAM) consensus clustering method, which is well-known for its robust performance in identifying the true underlying number of clusters within a data set, to cluster stage I LUAD patients using the expression profiles of COL11A1 and THBS2. The expression levels of COL11A1 and THBS2 were used to calculate the Euclidean distance between each sample and the center point of each cluster. After the number of clusters was evaluated from 2 to 10, we identified two subtypes (clusters) named S1 (low expression of COL11A1 and THBS2) and S2 (high expression of COL11A1 and THBS2). This clustering showed the clearest cut (between-cluster distance) and highest Average Silhouette Width (ASW)^14,15^, a popular cluster validation index to estimate the number of clusters within a data set (**Figures 3A** and **3B**).

**Figure 3.**
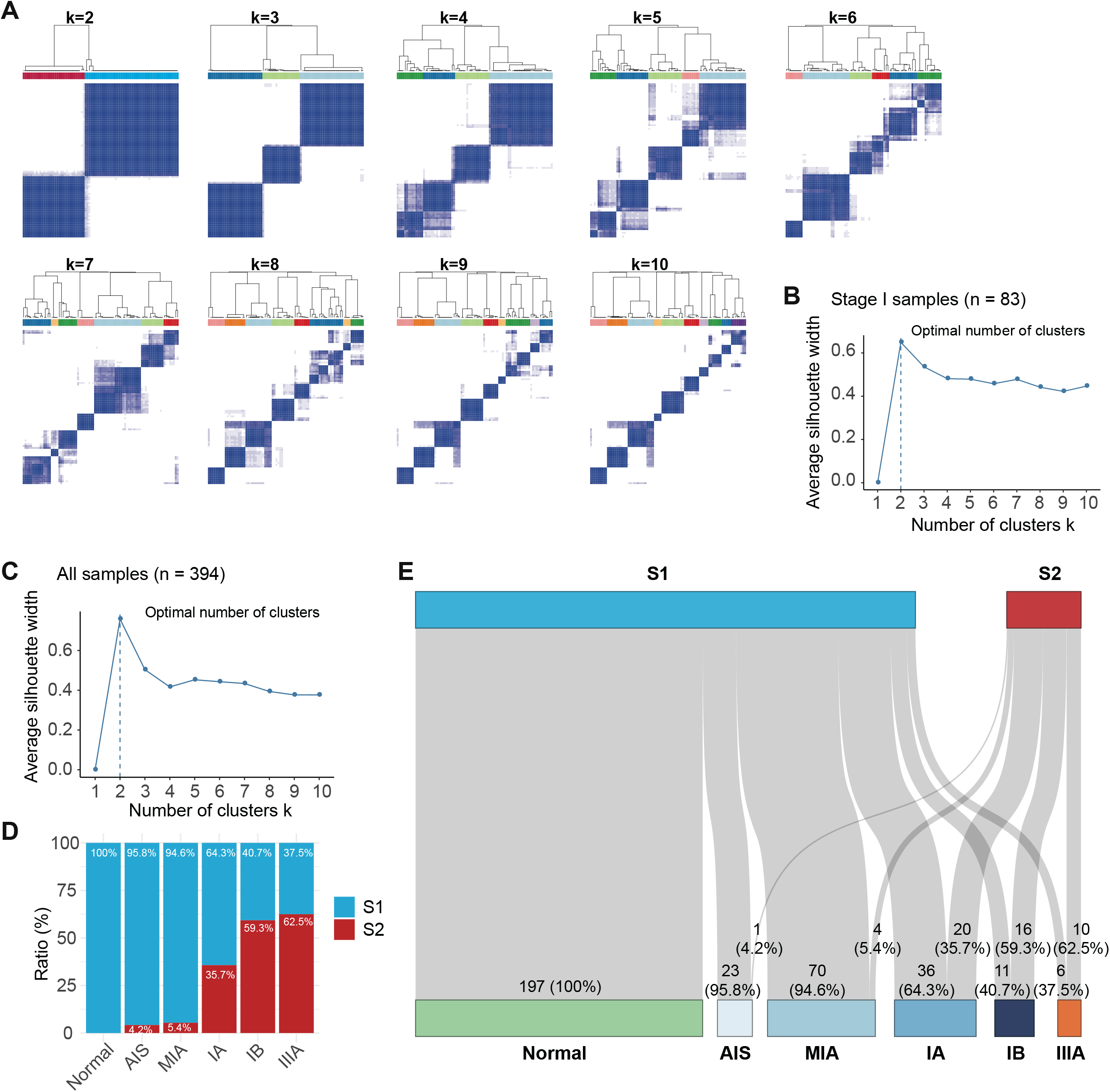
Molecular subtypes of stage I LUAD and all samples. (A) Consensus matrix for clusters ranging from 2 to 10 based on expression level of COL11A1 and THBS2. (B) The average silhouette width of 2-10 clusters for stage I LUAD samples, and the optimal number of clusters is two. (C) The average silhouette width of 2-10 clusters in the whole set of 394 samples, and the optimal number of clusters is two. (D and E) The distribution of samples classified as subtypes S1 and S2 for different pathological types of samples, from normal to stage IIIA.

To evaluate the molecular subtypes underlying the whole set of 394 samples including normal, AIS, MIA, and invasive LUAD, we performed PAM clustering using the expression profiles of COL11A1 and THBS2. Consistent with the clustering of stage I LUAD samples alone, the average silhouette width indicated that the optimal number of clusters was two (**Figure 3C**). It was interesting and gratifying to notice that 100% normal, 95.8% AIS, 94.6% MIA, 64.3% IA, 40.7% IB, and 37.5% IIIA were assigned to S1 (**Figures 3D** and **3E**). These results indicated that S1 was closer to AIS/MIA, and that as the disease stage progresses, more and more patients became S2-like.

### Distinct molecular differences between S1 and S2 subtypes

We extensively explored the differences in molecular characteristics between S1 and S2 subtypes within stage I LUAD, and included AIS/MIA as a control group in subsequent analyses. We identified seven genes with significantly different mutation frequency among AIS/MIA, S1, and S2 using Fisher’s exact test (**Figure 4A**). Four (EGFR, TP53, TTN, and CSMD3) of the seven genes were among the top 20 most frequently mutated genes in our data sets (**Figure S2A**). Except for EGFR and MGAM, the mutation frequency of the other five genes (TP53, TTN, CSMD3, DST, and FSCB) significantly increased in S2 over S1 (**Figure 4B**). Similarly, tumor mutation burden (TMB) gradually increased from AIS/MIA to S1 to S2 (**Figure 4C**). The same trend was also seen in APOBEC-related mutations, although only AIS/MIA vs S2 was significantly different (**Figure 4D**). These results indicated that S1 was genetically closer to AIS/MIA than S2.

**Figure 4.**
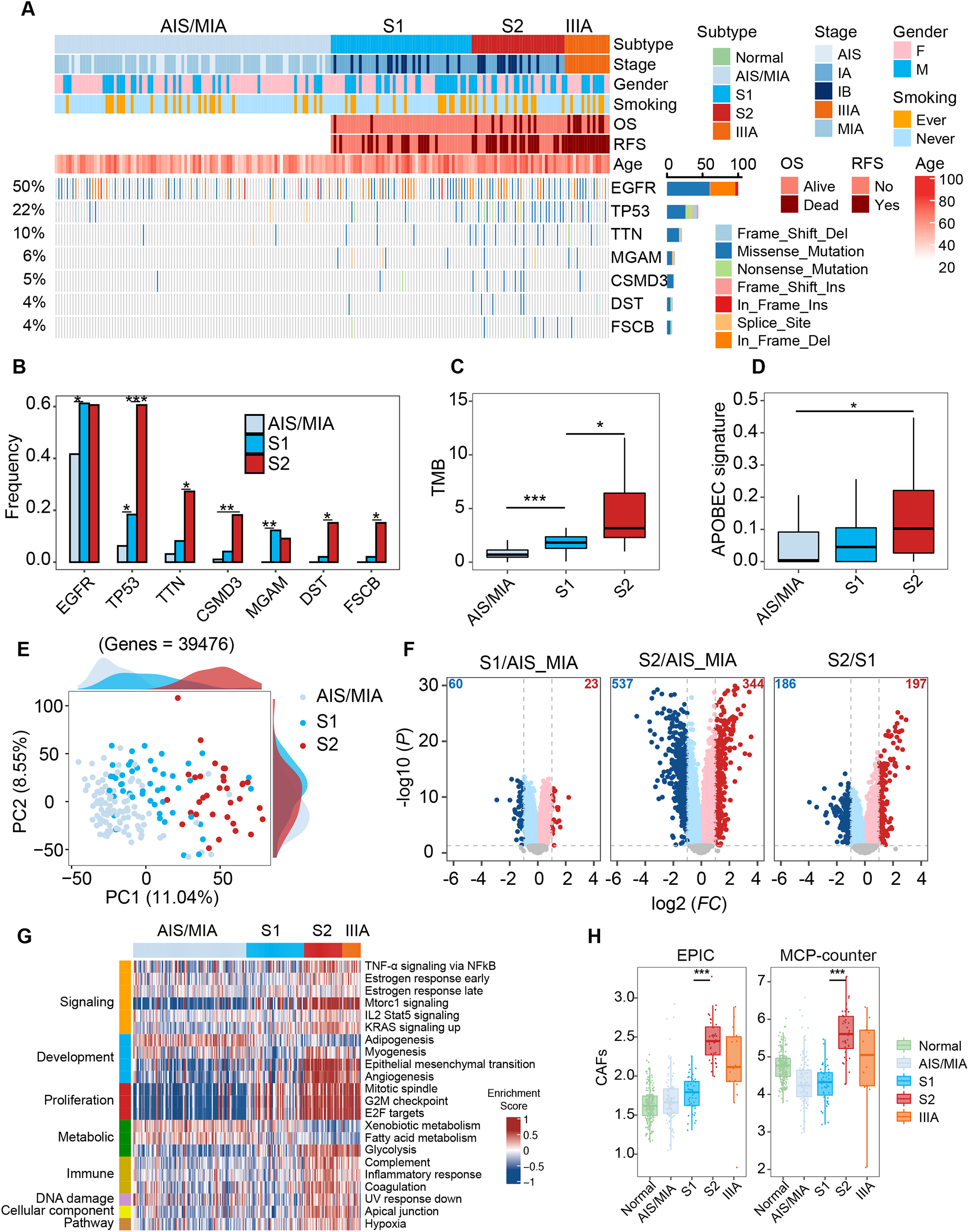
Molecular subtypes of stage I LUAD and associated distinct genomic and transcriptomic characteristics. (A) Classification of stage I LUAD into S1 and S2 subtypes. DMGs among AIS/MIA, S1, and S2 were shown from AIS/MIA to IIIA LUAD. (B) Gene mutation frequency of DMGs for AIS/MIA, S1, and S2. AIS/MIA had lower EGFR and MGAM mutation frequency than S1 and S2. Gene mutation frequency of TP53, TTN, CSMD3, DST, and FSCB increased significantly from AIS/MIA to S2. (C) Boxplot shows that S2 had higher TMB than S1 and AIS/MIA. (D) Boxplot shows that S2 had higher APOBEC-related mutation than AIS/MIA. (E) PCA of the expression profiles of 39,476 genes in AIS/MIA, S1, and S2. (F) Volcano plots show between-group differences in gene expression, S1 vs AIS/MIA, S2 vs AIS/MIA, and S2 vs S1. (G) Enrichment scores from the get set variation analysis of DEGs between AIS/MIA, S1, and S2. (H) Boxplots show CAF in different pathological stages, of which stage I was divided into S1 and S2. S2 had a higher CAF than S1. (^*^ *P* < 0.05, ^**^ *P* < 0.01, ^***^ *P* < 0.001).

Consistent with the trend of genomic characteristics, transcriptomic analysis also indicated that S1 was similar to AIS/MIA. PCA demonstrated that gene expression profiling of S1 was closer to AIS/MIA than S2 (**Figure 4E**). We further compared the expression profiles between AIS/MIA, S1 and S2, and identified 83 DEGs between AIS/MIA and S1, 881 DEGs between AIS/MIA and S2, and 383 DEGs between S1 and S2 (**Figure 4F**). Several pathways (ECM-receptor interaction, protein digestion and absorption, and focal adhesion) were enriched with most genes upregulated in S2 (**Figure S2B**). Meanwhile, the expression of all 15 FA pathway DEGs between AIS/MIA and invasive LUAD **(Figure 2E)** were also significantly altered between S1 and S2 (**Figure S2C**). We further explored cancer-associated biological functions of DEGs between AIS/MIA, S1, and S2 using Get Set Variation Analysis (GSVA). A total of 22 hallmarks of cancer were identified using hallmark gene sets from MSigDB (**Figure 4G**). The enrichment scores of these identified hallmarks showed continuous changes from AIS/MIA, S1, S2, to IIIA, indicating that S1 may be an intermediate biological stage during the development of AIS/MIA to S2 or IIIA.

Finally, we explored the differences in tumor microenvironment (TME) between AIS/MIA, S1, and S2. We analyzed the composition of TME using EPIC^17^ and MCP-counter^18^, two widely used software packages for such purposes. Associations between CAF and molecular subtypes were observed in that S2 with COL11A1 upregulation had more activated CAF than S1 (**Figure 4H**). Consistent with published research, COL11A1 in CAF was increased compared with normal fibroblasts in non-small cell lung cancer (NSCLC)^19^. CAF, one of the most abundant cell types in tumor tissues, was closely associated with promoting lung cancer development^20-22^. In fact, many clinical studies aimed at inhibiting the interplay between CAF and tumors are ongoing^22^. Meanwhile, several studies had suggested that CAF may promote cancer invasion by remodeling extracellular matrix (ECM)^22^. We also observed that CAF and the ECM-receptor interaction pathways were more active in S2 (**Figures 4H** and **S2B**), suggesting that CAF and ECM played important roles in LUAD progression. Therefore, patients with S2, which showed more activated CAF than S1 and AIS/MIA, may benefit from treatment aimed at preventing CAF activation.

### Distinct S1 and S2 subtypes were also observed with proteogenomic data

We subsequently reanalyzed the multi-omics data from the study of Gittelle *et al*.^23^ to explore differences in proteogenomic characteristics between S1 and S2 subtypes. Gene mutation and expression data were downloaded from Genomic Data Commons (GDC)^24^ and proteomics and phosphoproteomics data were downloaded from Clinical Proteomic Tumor Analysis Consortium (CPTAC)^25^. In this unique data set, PAM consensus clustering was also used to classify stage I LUAD based on the expression of COL11A1 and THBS2. Again, the optimal number of clusters was determined to be two after the average silhouette width was evaluated from 2 to 10 (**Figure S3A**). Therefore, PAM consensus clustering was performed to identify two subtypes based on the expression of COL11A1 and THBS2 of stage I patients. Consistent with what were observed in the FUSCC data set, S2 showed more mutation events, more dead or relapse events than S1 (**Figures 5A** and **5B**). The mutation frequency of TP53, RYR2, USH2A, KRAS, and XIRP2 was much higher in S2 than S1 (**Figure 5B**). Moreover, the event of copy number variations, such as amplification peaks, was less common in S1 than S2 (**Figure 5C**). Consistent with our FUSCC data set, the genomes of S1 were relatively simpler than S2.

**Figure 5.**
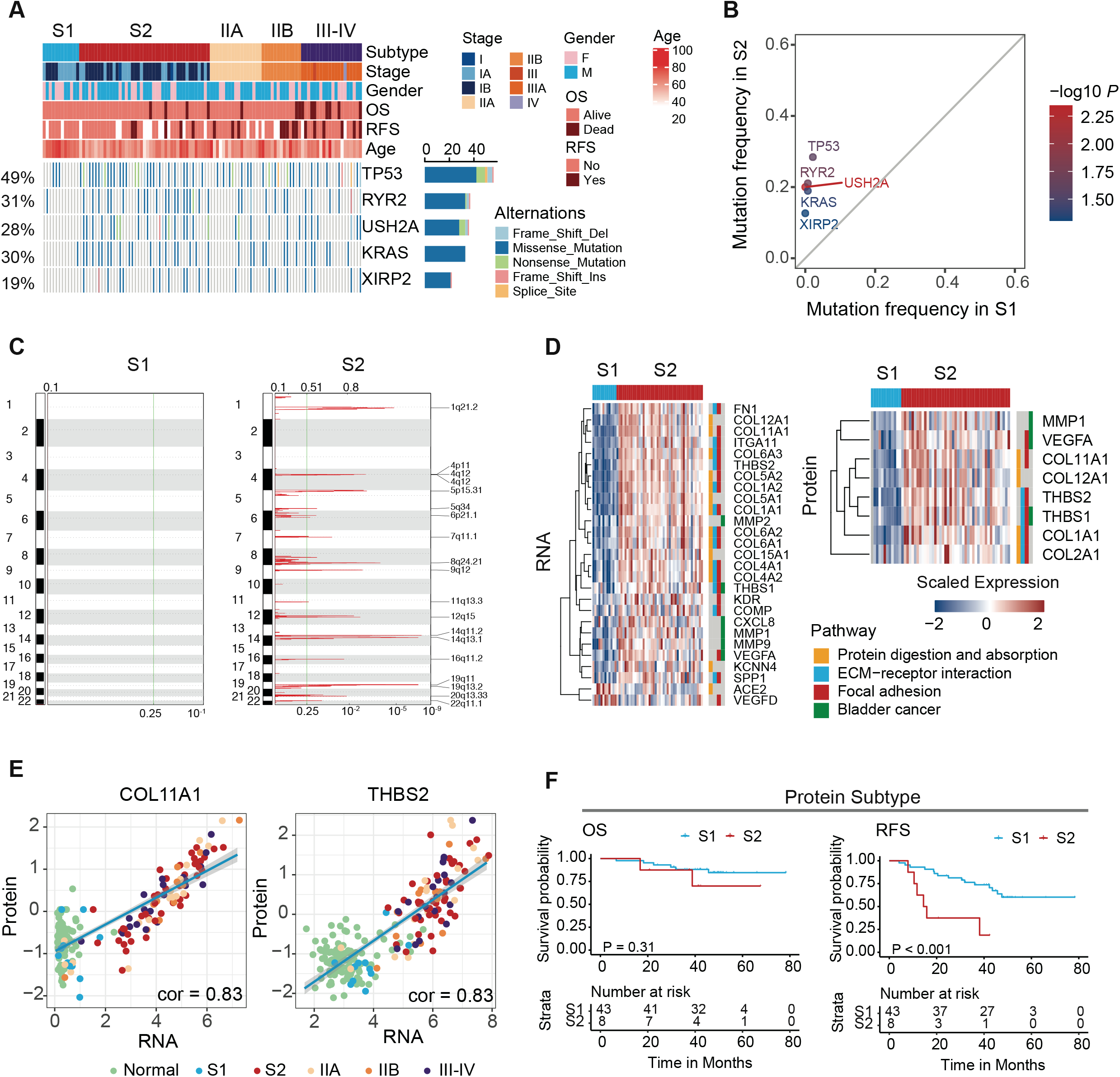
Proteogenomic relationships between S1 and S2 subtypes. (A) Classification of all samples into pathological clusters, of which stage I was divided into S1 and S2 subtypes. Oncoplot shows DMGs between S1 and S2. (B) S2 had a higher gene mutation frequency than S1. (C) Significant amplification peaks based on copy number profiling of S1 and S2. (D) Heatmaps show the DEGs and DEPs and the five pathways which were both enriched with DEGs and DEPs. (E) Scatterplots show the correlation between RNA and protein expression for COL11A1 and THBS2. (F) Clinical outcomes of S1 and S2 subtypes based on protein expression of COL11A1 and THBS2 in the Xu *et al*. data set. S1 had better RFS than S2.

Quantitative omics, including transcriptomics, proteomics, and phosphorylated proteomics analysis confirmed the distinct differences between S1 and S2. We performed differential expression analysis between S1 and S2 and identified 371 DEGs, 64 differentially expressed proteins (DEPs), and 121 differentially expressed phosphoproteins (DEPPs) (**Figure S3C**). To further explore biological functions associated with DEGs and DEPs, we performed KEGG pathway enrichment analysis. We found that DEGs and DEPs between S1 and S2 were both enriched in protein digestion and absorption, ECM−receptor interaction, focal adhesion, bladder cancer, and steroid hormone biosynthesis pathways (**Figure 5D**). Meanwhile, we also found that S2 showed more activated CAF than S1 (**Figure S3D**). Consistent with what we identified from our FUSCC data set, more activated CAFs and ECM−receptor interaction were also identified for S2 in the Gittelle *et al*. data set.

We observed a strong correlation between gene and protein expression levels for both COL11A1 and THBS2 (**Figures 5E, S3E** and **S3F**), indicating that protein expression, like gene expression, may also be used for molecular subtyping of stage I LUAD patients. To explore the relationship between clinical outcomes and molecular subtypes identified by consensus clustering based on the protein expression of COL11A1 and THBS2, we downloaded proteomics data and the corresponding clinical information from the study of Xu *et al*.^26^. We identified two subtypes closely associated with relapse-free survival (RFS) (*P* < 0.001), although the association with overall survival did not achieve statistical significance (*P* = 0.31, **Figure 5F**).

### AIS/MIA-like S1 subtype had better prognosis than AIS/MIA-diverging S2 subtype as validated with 11 external data sets

Similar molecular characteristics between S1 and AIS/MIA indicated that they may exhibit similarly excellent prognosis. In addition to our FUSCC data set, we downloaded 11 external published data sets containing both gene-expression data and prognosis information of stage I LUAD (**Table S1**). A total of 1,368 patients of stage I LUAD were involved in the 12 data sets. When each of the 12 data sets was subjected to the PAM analysis using the expression data of COL11A1 and THBS2, the optimal number of clusters for all data sets was found to be two (**Figure S4**), indicating the true number of underlying molecular subtypes of stage I LUAD. After subtyping each data set, we merged the 1,368 patients from the 12 data sets together and performed survival analysis (**Figure 6**). As we expected, survival analysis indicated that subtype S1 had better overall survival (**Figure 6A**, *P* < 0.0001) and relapse-free survival (**Figure 6D**, *P* < 0.001) than S2. Multivariate Cox analysis also indicated that the two-class subtyping was an independent prognostic variable (**Figures S5A** and **S5B**). We further compared the prognostic predictive performance of the two-class subtyping for patients within stage IA and IB separately. As a result, the OS and RFS of S1 were significantly better than S2 for stage IA patients (**Figures 6B** and **6E**, *P* < 0.001). However, the prognosis predictive performance of two-class subtyping for stage IB was not as good as in stage IA (**Figures 6C** and **6E**). Combining IA and S1 would help identify patients who may be truly at low risk (**Figures S5C** and **S5D**). These findings also suggested that the FA2 model had better prognostic stratification for early-stage IA.

**Figure 6.**
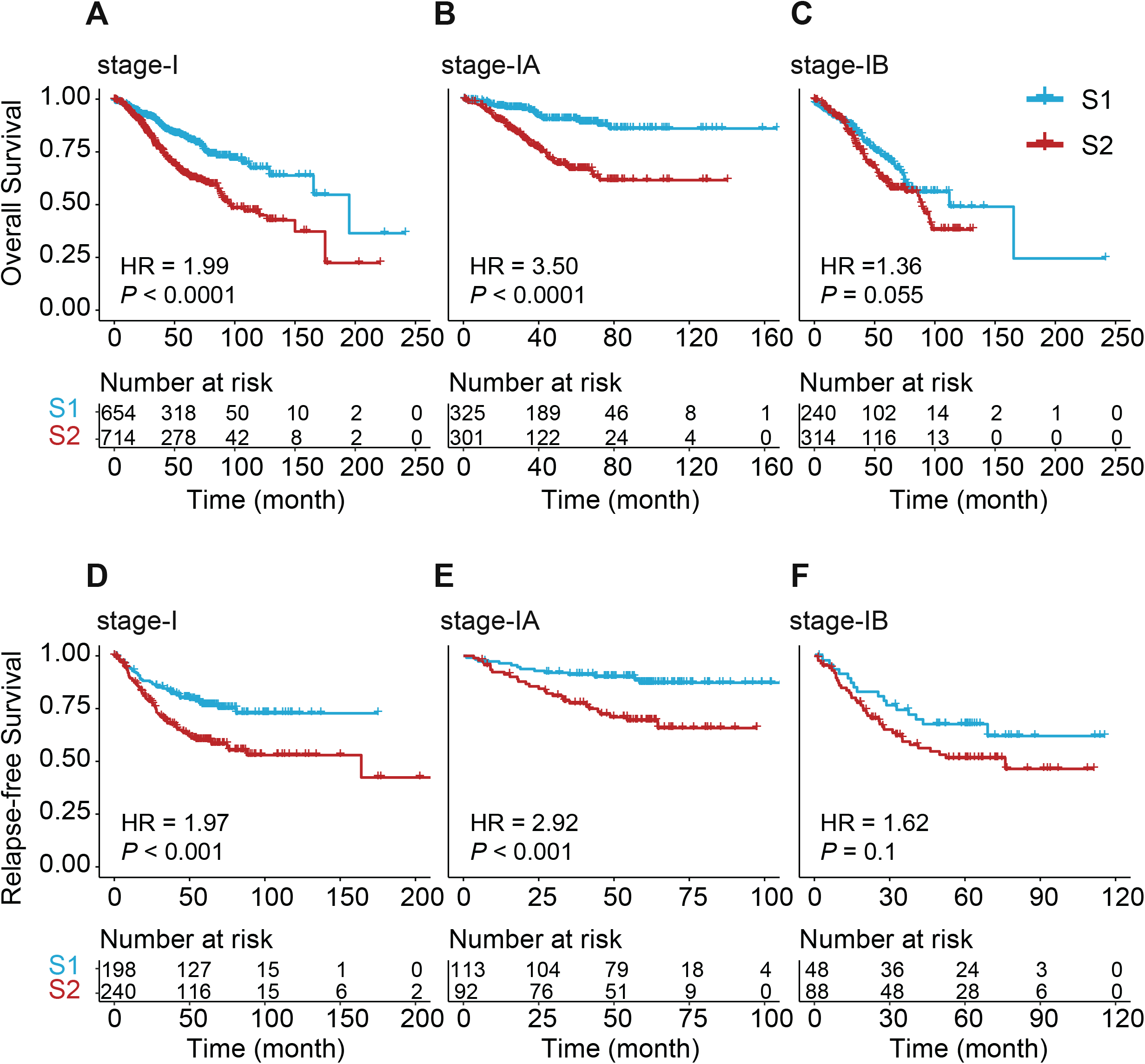
Survival analysis of FA2-based subtypes (S1 and S2) for 1,368 stage I patients from 12 cohorts. Overall survival (OS) for stage I (A), IA (B), and IB (C). S1 showed significantly better OS than S2 for stages I and IA (*P* < 0.01). Relapse-free survival (RFS) for stages I (D), IA (E), and IB (F). S1 showed significantly better RFS than S2 for stages I and IA (*P* < 0.01).

To comprehensively evaluate and compare the prognostic predictive power of the simple FA2 model involving COL11A1 and THBS2, we identified 42 published prognosis gene signatures of lung cancer through literature research and the review of Tang *et al*.^7,9,27^, with mean and median number of genes of 65 and 42, respectively (**Table S3**). PAM consensus clustering was performed using the expression profiles of the 42 gene signatures to identify two subtypes for the stage I patients within each of the 12 data sets. To facilitate fair comparisons between different signatures (models), the optimal number of clusters was chosen to be two for all models. We merged the 1,368 patients from the 12 data sets together after we finished subtype classification for each data set. AUC of time-dependent receiver-operating characteristics (ROC) curve and concordance index (C-index) were used to evaluate the performance of the 42 literature signatures plus the FA2 signature with the 1,368 patients. The 43 models were ranked by the mean of AUC and C-index. As a result, the FA2 model, with the least number of genes, ranked top three (OS, **Figure 7A**) and top five (RFS, **Figure 7B**) in prognostic predictive power for stage IA patients only. For all stage I patients, the FA2 model ranked top five (OS, **Figure S6A**) and top 15 (RFS, **Figure S6B**). These results further suggested that FA2 genes were biologically important and had a good predictive performance for prognosis of stage I patients, especially stage IA patients.

**Figure 7.**
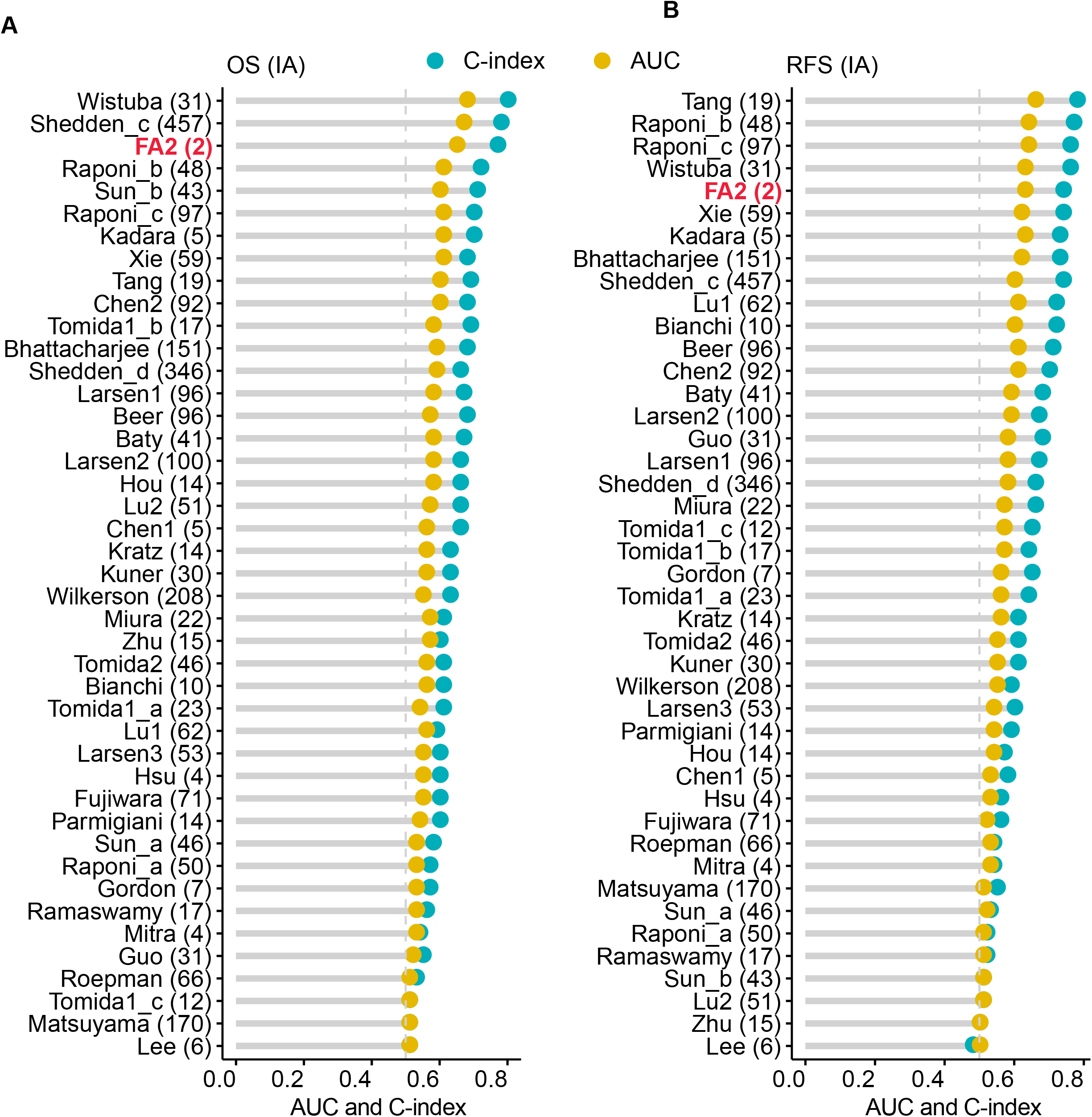
Comparison of performances of 43 prognostic predictive models for stage IA patients. Models were ranked by the mean of AUC and C-index for five-year overall survival (OS, A) and relapse-free survival (RFS, B). The number within the paratheses following the name of a model refers to the number of genes used in the model. The mean and median number of genes over the 42 published models is 65 and 42, respectively. The FA2 model with the least number of genes (two) ranked top three and top five for OS and RFS, respectively.

## DISCUSSION

Changes in molecular characteristics from pre-invasive AIS/MIA to invasive LUAD may provide us with insights for the accurate classification of stage I LUAD with divergent prognosis. Many studies on constructing models for risk stratification of stage I LUAD based on gene expression have been reported^11,12,27^. It is usually a straightforward choice to obtain gene features of “high” prognostic prediction performance through training with prognosis as the endpoint, but this approach is prone to over-fitting^10^. Different from previous studies, we identified two genes (COL11A1 and THBS2) in the FA pathway based mainly on somatic alterations and gene-expression alterations between AIS/MIA and invasive LUAD. In the process of selecting features and conducting molecular subtyping, we did not perform any training with knowledge of a patient’s prognosis and thus effectively avoided overfitting, as can be seen from the performance validation results with 11 external data sets.

The two genes played important roles in the progression of lung adenocarcinoma. COL11A1 showed higher mutation frequency in invasive LUAD than AIS/MIA (**Figure 2C**). Meanwhile, the expression of COL11A1 was almost undetectable in normal and AIS/MIA, but started to increase dramatically in stage IA LUAD (**Figure 2I**). These results indicated that COL11A1 may promote AIS/MIA progression. Many studies have also demonstrated that COL11A1 plays an important role in tumor progression including NSCLC^28-31^. Similarly, there was no significant difference in the expression of THBS2 between normal and AIS/MIA, whereas there was a steady increase from IA to IB and IIIA LUAD (**Figure 2I**). The evolving mutation or expression characteristics of COL11A1 and THBS2 indicated their close association with the invasiveness of LUAD. Indeed, FA2 consisting of COL11A1 and THBS2 helped identify two molecular subtypes S1 and S2 with different degrees of invasion in stage I LUAD.

The consistency and robustness of genomic, transcriptomic, and proteomic differences between S1 and S2 demonstrated that the two subtypes of stage I LUAD were biologically and clinically relevant. In our FUSCC data set, the genomic and transcriptomic characteristics of S1 were similar to AIS/MIA, and the prognosis of S1 was closer to AIS/MIA. Furthermore, the high correlation between gene and protein expression for COL11A1 and THBS2 indicated that protein expression data may also be used for molecular classification. We performed consensus clustering using protein expression of COL11A1 and THBS2 to divide stage I LUAD into S1 and S2 using a publicly available multi-omics data set. The significant differences in RFS between S1 and S2 further suggested the prospect of clinical applications of COL11A1 and THBS2.

The differences in molecular characteristics between S1 and S2 may have potential therapeutic implications. The poor clinical outcomes and high TMB of S2 may provide hint for clinical decision-making. At present, the guidance for clinical adjuvant therapy is still based on pathological staging. What is more, there is no obvious evidence that patients with stage I LUAD can benefit from adjuvant therapy, so that most stage I patients do not undergo adjuvant therapy systematically after surgery^5,6^. According to our analysis, the classification of stage I LUAD into S1 and S2 was better than IA and IB in prognostic predictive performance (**Figure S5**). Stage I LUAD patients who were classified as S2 had a higher risk of recurrence or death. At the same time, S2 had higher TMB than S1. Many studies have shown that patients with high TMB may benefit from immune checkpoint inhibitors^32^. Thus, our results indicated that S2 patients may benefit from receiving adjuvant therapy, such as immunotherapy. Meanwhile, a high score of CAF in S2 may suggest possible treatment. Many studies found that TME played an important role in the development of tumor invasion^33,34^. CAF, an important component in the TME, is distributed among tumor cells to provide a beneficial tumor stroma^22^. It was reported that CAF promoted cancer invasion by remodeling ECM^22,35^. Coincidentally, CAF level was almost not changed among paired normal, AIS/MIA and S1, but there was obvious activation in S2 (**Figures 4H** and **S3D**). Furthermore, the ECM-receptor interaction pathway was more active in S2 than in S1 (**Figures S2B** and **5D**). These results indicated that CAF may promote early LUAD invasion by remodeling ECM. In fact, several clinical trials of targeted therapeutic for the interaction between CAF and tumor have been initiated^22^. Our results suggested that S2 patients may benefit from drugs that attenuate CAF activation.

Compared with published models, the FA2 model proposed in our study consisted of the least number of genes (two), but performed better than most published gene signatures (**Figures 7** and **S6**). Furthermore, the prognostic predictive power of FA2 was better than pathological staging and FA2 was an independent prognosis predictor (**Figures 6** and **S5**). These results suggested that FA2 had the potential to complement pathological staging. However, there were some limitations in the evaluation of the performance of FA2 compared to published gene signatures. The best evaluation approach would be combining the published gene signatures with the corresponding classification methods, and then evaluating the performance of each published signature-classification method pair. However, it is very difficult if not impossible for us to completely replicate the published method^7^. Therefore, we compared the prognostic predictive performance of published gene signatures using the same widely adopted unsupervised clustering algorithm of PAM for its ability of finding the optimal number of clusters underlying a data set.

In conclusion, we applied PAM consensus clustering with COL11A1 and THBS2 expression to classify stage I LUAD into S1 (AIS/MIA-like) and S2 (CAF-rich) subtypes, which showed clear differences in multi-omics, tumor microenvironment, and clinical outcomes. The molecular classification of stage IA and stage I LUAD showed good prognosis predictive performance, which may provide more precise management of these patients in clinical practice. S2 (CAF-rich) with higher TMB and CAF may benefit from adjuvant therapies, such as immunotherapy and CAF suppression therapy. However, prospective studies and functional or mechanistic experiments need to be completed to further verify our conclusions. Nevertheless, our simple and robust FA2 model may serve as a foundation for reliable identification of high-risk stage I LUAD patients for more intensive post-surgery treatment.

## METHODS

### Patients

As described in our previous article^13^, we collected tumor-normal matched samples from a total of 197 patients during surgery, including AIS, MIA, IA, IB, and IIIA. No patient received neoadjuvant therapy before surgery. In this study we added further information on the prognosis of these patients at follow-up. RFS and OS time was recorded according to clinical or telephone follow-up. This study has been approved by the research ethics review committee of Fudan University Shanghai Cancer Center (FUSCC) Institutional Review Board (No. 090977-1).

### RNA-seq data analysis

Hisat2-StringTie pipeline was used to obtain expression profiles from raw FASTA data^36^. Trimmomatic (v0.36) was used to remove adapters in the raw RNA-seq reads^37^. The quality of raw RNA-seq reads was assessed through FastQC (v0.11.5). FastQ Screen (v0.11.0) was used to evaluate whether there was contamination from other species in RNA-seq reads. We used Hisat2 (v2.0.5) to align reads to the human reference genome (GRCh38, release-84), which was downloaded from GDC^36^. The reads aligned to the human reference genome were assembled by StringTie (v1.3.3) and annotated as transcripts or genes by genome annotation file (gencode.v22.annotation.gtf)^36^. Finally, Fragments Per Kilobase of exon model per Million mapped fragments (FPKM) was used to measure gene expression.

### Mutation profiling

As described in the previous study^13^, the gene mutation data were generated through whole-exome sequencing (WES). In this study, we continued to use the results of the previous analysis. The TMB- and APOBEC-related mutation data also came from the previous study and can be downloaded from the supplementary information (https://doi.org/10.1038/s41467019-13460-3).

### Processing of publicly available data sets

Data collection was conducted from October 2020 to March 2021. All gene mutation data of TCGA and Gillette *et al*.^23^ were downloaded from GDC^24^. All gene expression microarray data and corresponding clinical phenotypes were obtained from Gene Expression Omnibus (GEO) (https://www.ncbi.nlm.nih.gov/geo/). Gene symbols were used to represent genes from different platforms. If there were multiple probes corresponding to the same gene symbol, the one with the highest signal intensity was used to represent the expression level of the corresponding gene. RNA-seq gene expression data sets (TCGA and Gillette *et al*.^23^) and corresponding clinical phenotypes were downloaded from GDC. The proteomic data of the two studies used in our analysis were obtained as follows. Normalized protein and phosphorylated protein expression data of Gillette et al. were downloaded from Clinical Proteomic Tumor Analysis Consortium (CPTAC)^25^. Normalized protein expression data of Xu *et al*.^26^ were downloaded as an attachment table of the article. As shown in **Table S1**, our study used 14 published data sets including two genomic data sets, 11 microarray gene expression data sets, two RNA-seq transcriptomics data sets, two proteomics data sets, and one phosphoproteomics data set. Summary of the published data sets was shown in **Table S2**.

### Differential gene expression and mutation analysis

R package limma (v3.42.2) was used to perform differential expression analysis between normal, pre-invasive, and invasive LUAD, or between S1 and S2 subtypes^38^. The commonly used cutoffs (*P* < 0.05, |log2(fold change)| >= 1) were used to identify differentially expressed genes^39^. Fisher’s exact (*P* < 0.05) was used to identify differentially mutated genes between pre-invasive and invasive LUAD or between S1 and S2 subtypes.

### KEGG and ssGSEA analysis

R package clusterProfiler (v3.14.3) was used to perform KEGG pathway enrichment analysis^40^. Focal adhesion pathway genes and hallmark gene sets were downloaded from MSigDB. Pathways significantly enriched with genes in an input set were identified by adjusted *P*-value (*P* < 0.05). R package GSVA^41^(v1.34.0) with default gsva method was used to estimate gene-set enrichment score.

### Partition Around Medoids (PAM)

Unsupervised clustering using the partition around medoids (PAM) cluster algorithm and Euclidean distance was performed through R package ConsensusClusterPlus (v1.50.0)^42^. The two genes (COL11A1 and THBS2) in the FA pathway, which were commonly identified both as DEGs or DMGs between AIS/MIA and invasive LUAD, were used as features for consensus clustering. R package factoextra (v1.0.7) was used to count average silhouette width^15^ and choose the optimal number of clusters in a data set. The consensus matrix with K = 2 was selected for further analysis after the number of clusters was evaluated from 2 to 10.

### Assessment of tumor microenvironment

EPIC^17^ and MCP-counter^18^ were used to identify the composition and density of cells based on gene expression of each sample. R package immunedeconv (v2.0.3)^43^ was used to perform EPIC and MCP-counter functions.

### Statistical analysis

All statistical analysis was performed with R (v3.6.3). Statistical tests included t-test, Fisher’s exact test, and Pearson correlation. R package survival (v3.1-8) and survminer (0.4.8) were used to perform survival and Cox analysis. Kaplan-Meier survival analysis combined with log-rank test was used for overall survival (OS) and relapse-free survival (RFS) analysis. R package ComplexHeatmap (v2.2.0) was used to draw heatmaps^44^. Principal component analysis (PCA) was performed using R package stats (v3.6.3). Oncoplot and lollipop plots were performed with maftools (v2.6.05)^45^. Amplification peaks were identified by GISTIC2.0. Boxplots and scatter plots were drawn with R package ggpubr (v0.4.0) and ggplot2 (v3.3.3).

## Data Availability

All data produced in the present study are available upon reasonable request to the authors

## ACKNOWLEDGEMENTS

This study was supported in part by the National Key R&D Program of China (2018YFE0201603), Shanghai Municipal Science and Technology Major Project (2017SHZDZX01), and the National Natural Science Foundation of China (31720103909). Fig. 1 was created with Biorender.com.

## AUTHOR CONTRIBUTIONS

YZheng, HC and JS conceived the study. JS, HJ and JY analyzed the data. YZhao collected and interpreted clinical data. NZ, LR, QC, and YY participated in the verification and interpretation of the data. JS and HJ drafted the manuscript. LS, HC and YZheng revised the manuscript and supervised the work. All authors reviewed and approved the manuscript.

## FIGURE LEGENDS

**Figure S1**. Transcriptomic and genomic alterations between AIS/MIA and invasive LUAD. (A) Comparison of gene mutation frequency between AIS and MIA. Color bar shows -log10 (*P*). (B) Volcano plot shows that there were almost no differences in gene expression between AIS and MIA. (C and D) Oncoplots show the somatic alterations of DMGs between AIS/MIA and invasive LUAD. (E) KEGG pathways enriched with DEGs between AIS/MIA and LUAD. (F) KEGG pathways enriched with DMGs between AIS/MIA and LUAD.

**Figure S2**. Genomic and transcriptomic alterations from AIS/MIA to S2 in the FUSCC data set. (A) Oncoplot shows mutation distribution of top 20 genes and six kinds of base conversion distribution ratio in AIS/MIA, S1, and S2. (B) The top five pathways enriched with DEGs between S1 and S2 were shown from normal to IIIA LUAD. (C) Boxplots show expression of 15 DEGs in the FA pathway from normal to stage IIIA.

**Figure S3**. The differences between S1 and S2 in transcriptomics, proteomics, and phosphorylated proteomics. (A) The average silhouette width of 2-10 clusters in stage I LUAD samples and the optimal number of clusters is two. (B) Classification of all samples into pathological clusters, of which stage I was divided into S1 and S2 subtypes. Heatmaps show differentially expressed genes (DEGs), differentially expressed proteins (DEPs), and differentially expressed phosphorylated proteins (DEPPs) between S1 and S2. (C) Volcano plots show DEGs, DEPs, and DEPPs between S1 and S2. Dark red and dark blue represent up-regulated and down-regulated, respectively. (D) Boxplots show CAF in different pathological stages, of which stage I was divided into S1 and S2. S2 had a higher CAF than S1. (E) Boxplots show expression of COL11A1 and THBS2 in normal, S1, S2, IIA, IIB, and III-IV LUAD. (F) Boxplots show protein expression of COL11A1 and THBS2 in normal, S1, S2, IIA, IIB, and III-IV LUAD. (^*^ *P* < 0.05, ^**^ *P* < 0.01, ^***^ *P* < 0.001, ^****^ *P* < 0.0001).

**Figure S4**. Determination of the optimal number of clusters in 11 published data sets. The Y-axis shows the average silhouette width of 2-10 clusters (K). The optimal number of clusters is two for all 11 data sets.

**Figure S5**. Multivariate Cox and survival analysis for five-year overall survival (OS) and relapse-free survival (RFS). The forest diagrams show the hazard ratio (HR) value and *P*-value of OS (A) and RFS (B) for subtype, pathological stage, age, gender, and smoking calculated through multivariate Cox analysis. OS (C) and RFS (D) for IA-S1, IA-S2, IB-S1, and IB-S2.

**Figure S6**. Comparison of performances of 43 prognostic predictive models for stage I patients. Models were ranked by the mean of AUC and C-index for five-year overall survival (OS, A) and relapse-free survival (RFS, B). The number within the paratheses following the name of a model refers to the number of genes used in the model. The mean and median number of genes over the 42 published models is 65 and 42, respectively. The FA2 model with the least number of genes (two) ranked top five and top 15 for OS and RFS, respectively.

