## supplementary files for "Poor prognosis of stage I lung adenocarcinoma patients determined by elevated expression over pre/minimally invasive status of COL11A1 and THBS2 in the focal adhesion pathway"

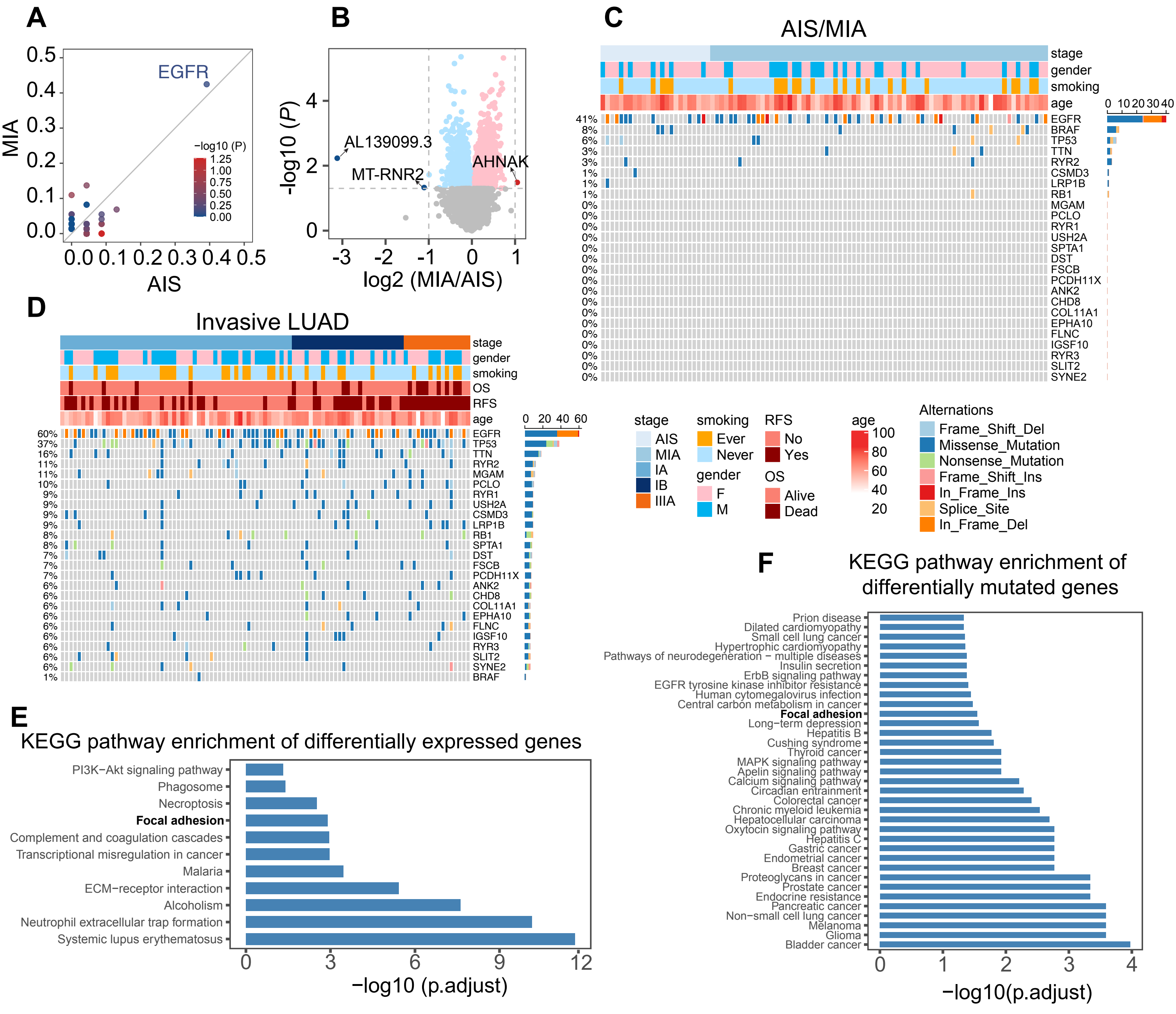

A
MIA
AIS
EGFR
−log10 (P)
B
-log10 (P)
log2 (MIA/AIS)
AL139099.3
MT-RNR2
AHNAK
C
AIS/MIA
stage
gender
smoking
age
EGFR
BRAF
TP53
TTN
RYR2
CSMD3
LRP1B
RB1
MGAM
PCLO
RYR1
USH2A
SPTA1
DST
FSCB
PCDH11X
ANK2
CHD8
COL11A1
EPHA10
FLNC
IGSF10
RYR3
SLIT2
SYNE2
D
Invasive LUAD
stage
gender
smoking
OS
RFS
age
EGFR
TP53
TTN
RYR2
MGAM
PCLO
RYR1
USH2A
CSMD3
LRP1B
RB1
SPTA1
DST
FSCB
PCDH11X
ANK2
CHD8
COL11A1
EPHA10
FLNC
IGSF10
RYR3
SLIT2
SYNE2
BRAF
stage
AIS
MIA
IA
IB
IIIA
smoking
Ever
Never
gender
F
M
RFS
No
Yes
OS
Alive
Dead
age
Alternations
Frame_Shift_Del
Missense_Mutation
Nonsense_Mutation
Frame_Shift_Ins
In_Frame_Ins
Splice_Site
In_Frame_Del
E
KEGG pathway enrichment of differentially expressed genes
PI3K−Akt signaling pathway
Phagosome
Necroptosis
Focal adhesion
Complement and coagulation cascades
Transcriptional misregulation in cancer
Malaria
ECM−receptor interaction
Alcoholism
Neutrophil extracellular trap formation
Systemic lupus erythematosus
−log10 (p.adjust)
F
KEGG pathway enrichment of differentially mutated genes
Prion disease
Dilated cardiomyopathy
Small cell lung cancer
Hypertrophic cardiomyopathy
Pathways of neurodegeneration − multiple diseases
Insulin secretion
ErbB signaling pathway
EGFR tyrosine kinase inhibitor resistance
Human cytomegalovirus infection
Central carbon metabolism in cancer
Focal adhesion
Long−term depression
Hepatitis B
Cushing syndrome
Thyroid cancer
MAPK signaling pathway
Apelin signaling pathway
Calcium signaling pathway
Circadian entrainment
Colorectal cancer
Chronic myeloid leukemia
Hepatocellular carcinoma
Oxytocin signaling pathway
Hepatitis C
Gastric cancer
Endometrial cancer
Breast cancer
Proteoglycans in cancer
Prostate cancer
Endocrine resistance
Pancreatic cancer
Non−small cell lung cancer
Melanoma
Glioma
Bladder cancer
−log10(p.adjust)


FigureS1

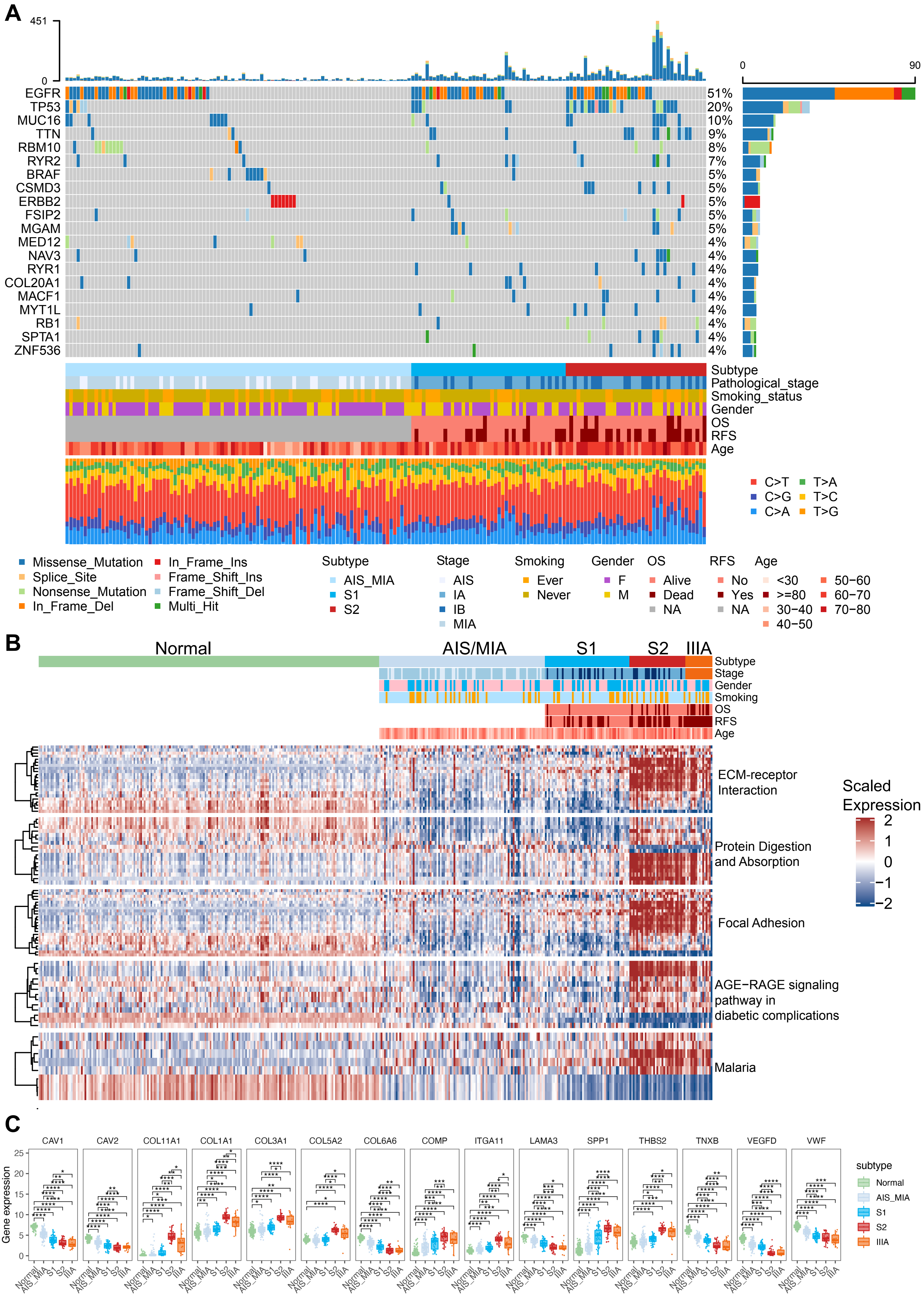

A
451
0
EGFR 51%
TP53 20%
MUC16 10%
TTN 9%
RBM10 8%
RYR2 7%
BRAF 5%
CSMD3 5%
ERBB2 5%
FSIP2 5%
MGAM 5%
MED12 4%
NAV3 4%
RYR1 4%
COL20A1 4%
MACF1 4%
MYT1L 4%
RB1 4%
SPTA1 4%
ZNF536 4%
0
90
Subtype
Pathological_stage
Smoking_status
Gender
OS
RFS
Age
C>T
T>A
C>G
T>C
C>A
T>G
Missense_Mutation
Splice_Site
Nonsense_Mutation
In_Frame_Del
In_Frame_Ins
Frame_Shift_Ins
Frame_Shift_Del
Multi_Hit
Subtype
AIS_MIA
S1
S2
Stage
AIS
IA
IB
MIA
Smoking
Ever
Never
Gender
F
M
OS
Alive
Dead
NA
RFS
No
Yes
NA
Age
<30
>=80
30−40
40−50
50−60
60−70
70−80
B
Normal
AIS/MIA
S1
S2
IIIA
Subtype
Stage
Gender
Smoking
OS
RFS
Age
ECM-receptor Interaction
Protein Digestion and Absorption
Focal Adhesion
AGE−RAGE signaling pathway in diabetic complications
Malaria
Scaled Expression
2
1
0
−1
−2
C
CAV1
CAV2
COL11A1
COL1A1
COL3A1
COL5A2
COL6A6
COMP
ITGA11
LAMA3
SPP1
THBS2
TNXB
VEGFD
VWF
Gene expression
25
20
15
10
5
0
Normal
AIS_MIA
S1
S2
IIIA
subtype
Normal
AIS_MIA
S1
S2
IIIA


FigureS2

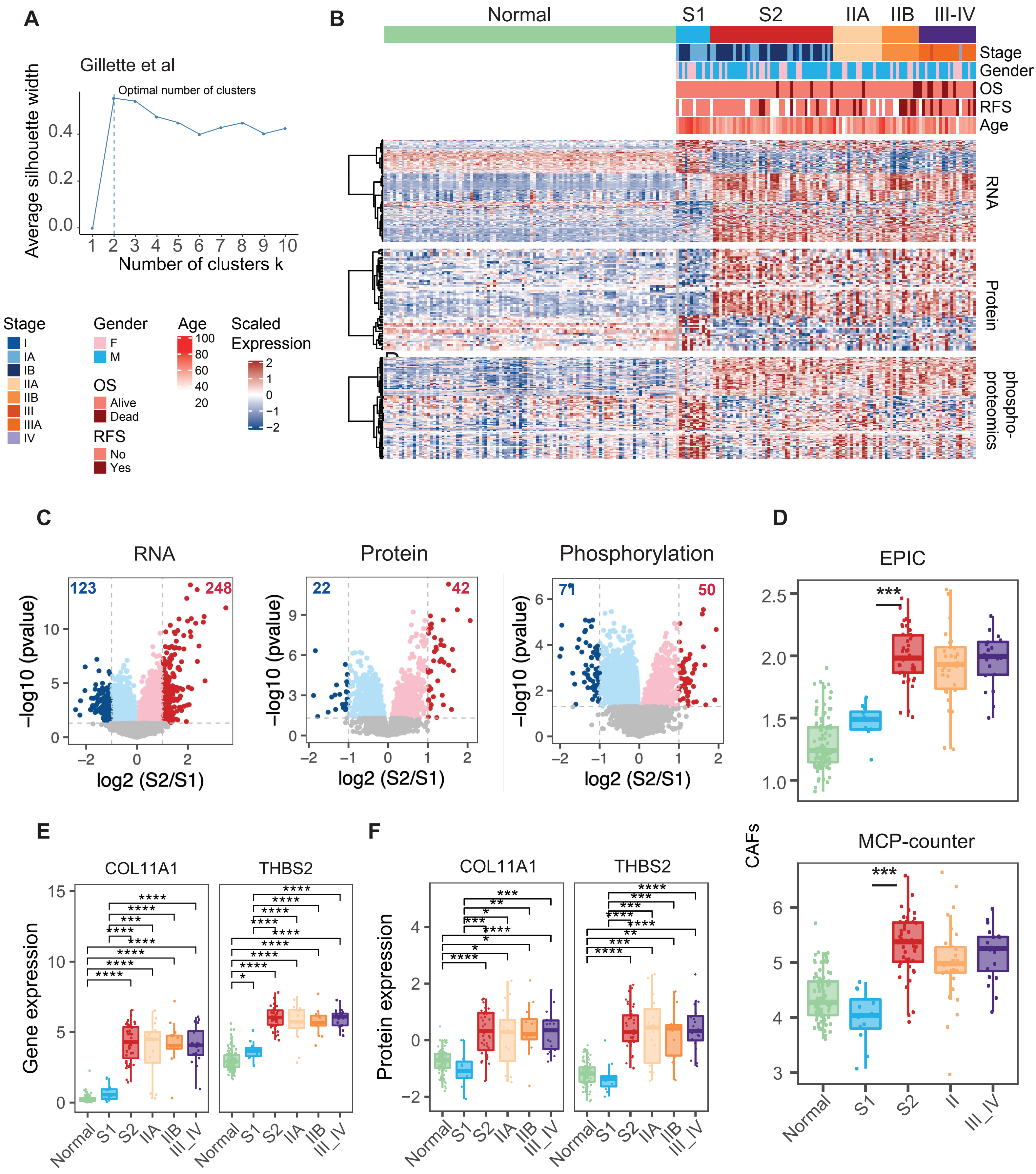

A
Gillette et al
Optimal number of clusters
Average silhouette width
Number of clusters k
B
Normal
S1
S2
IIA
IIB
III-IV
Stage
Gender
OS
RFS
Age
RNA
Protein
phospho-proteomics
Stage
I
IA
IB
IIA
IIB
III
IIIA
IV
Gender
F
M
OS
Alive
Dead
RFS
No
Yes
Age
Scaled Expression
C
RNA
Protein
Phosphorylation
−log10 (pvalue)
log2 (S2/S1)
D
EPIC
MCP-counter
CAFs
E
COL11A1
THBS2
Gene expression
F
Protein expression
Normal
S1
S2
IIA
IIB
III_IV


FigureS3

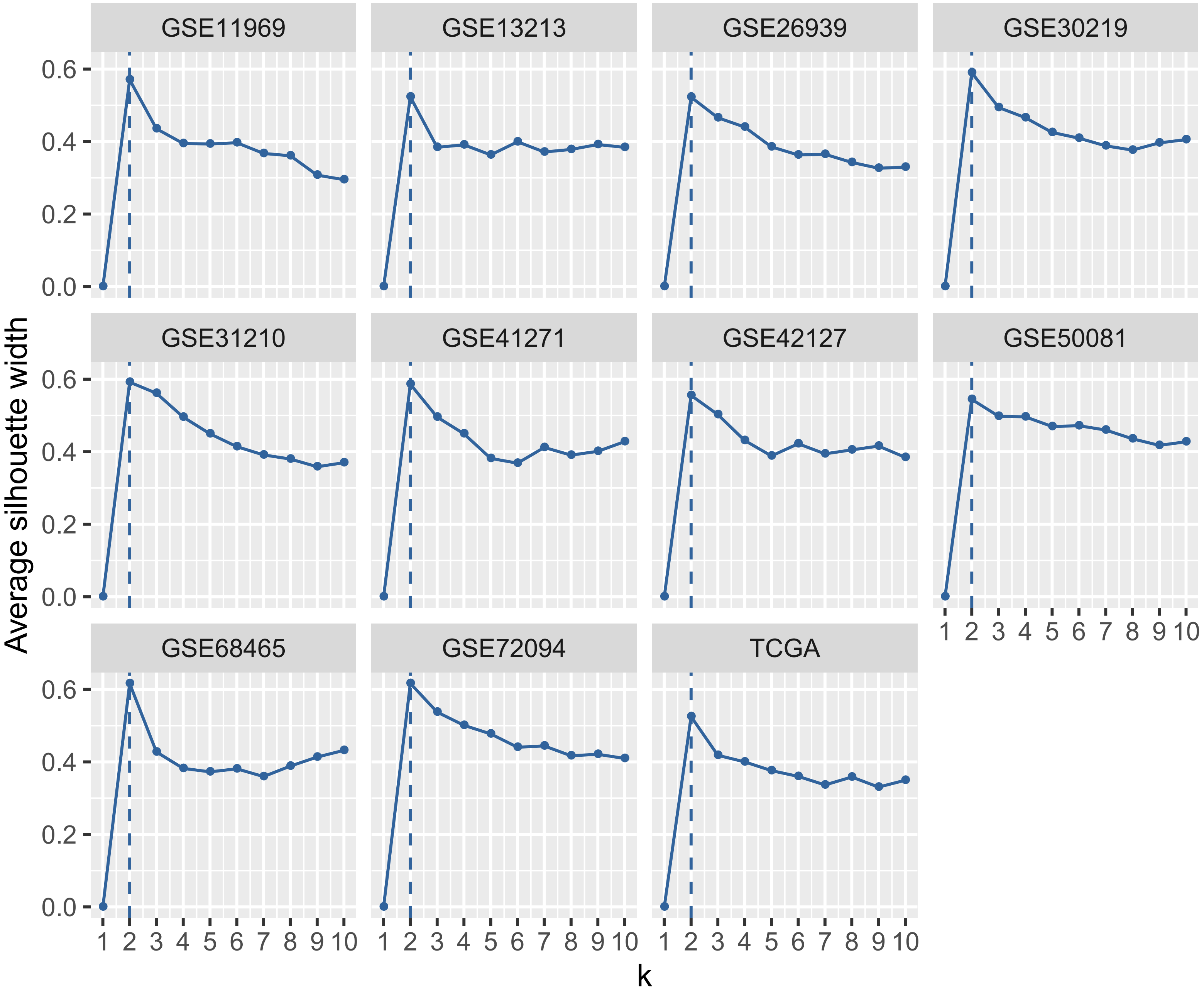

GSE11969
GSE13213
GSE26939
GSE30219
GSE31210
GSE41271
GSE42127
GSE50081
GSE68465
GSE72094
TCGA
Average silhouette width
0.0
0.2
0.4
0.6
1 2 3 4 5 6 7 8 9 10
k


FigureS4

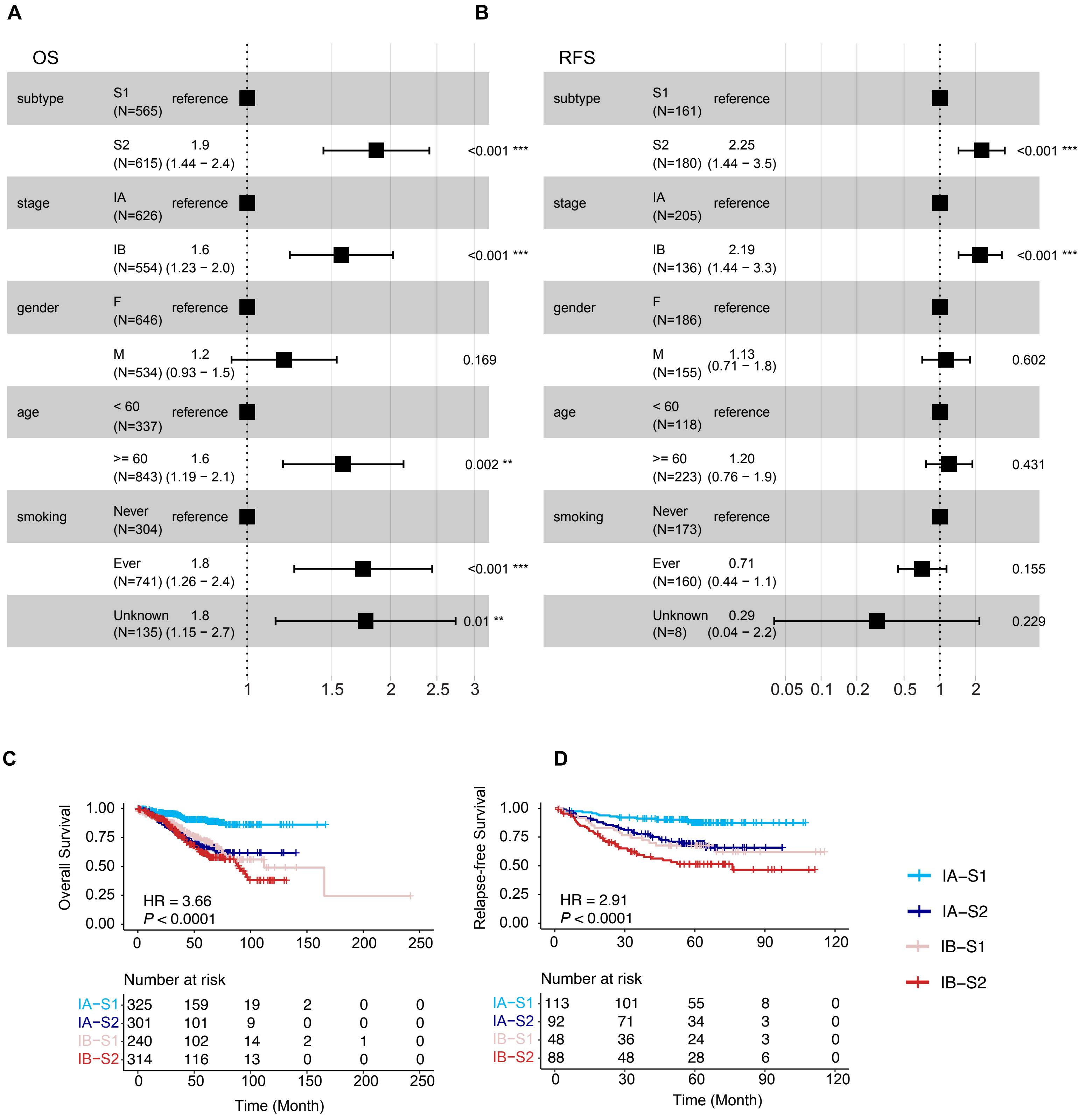
A
OS
subtype
S1 (N=565) reference
S2 (N=615) 1.9 (1.44 − 2.4) <0.001 ***
stage
IA (N=626) reference
IB (N=554) 1.6 (1.23 − 2.0) <0.001 ***
gender
F (N=646) reference
M (N=534) 1.2 (0.93 − 1.5) 0.169
age
< 60 (N=337) reference
>= 60 (N=843) 1.6 (1.19 − 2.1) 0.002 **
smoking
Never (N=304) reference
Ever (N=741) 1.8 (1.26 − 2.4) <0.001 ***
Unknown (N=135) 1.8 (1.15 − 2.7) 0.01 **
1 1.5 2 2.5 3
B
RFS
subtype
S1 (N=161) reference
S2 (N=180) 2.25 (1.44 − 3.5) <0.001 ***
stage
IA (N=205) reference
IB (N=136) 2.19 (1.44 − 3.3) <0.001 ***
gender
F (N=186) reference
M (N=155) 1.13 (0.71 − 1.8) 0.602
age
< 60 (N=118) reference
>= 60 (N=223) 1.20 (0.76 − 1.9) 0.431
smoking
Never (N=173) reference
Ever (N=160) 0.71 (0.44 − 1.1) 0.155
Unknown (N=8) 0.29 (0.04 − 2.2) 0.229
0.05 0.1 0.2 0.5 1 2
C
Overall Survival
1.00 0.75 0.50 0.25 0.00
HR = 3.66
P < 0.0001
0 50 100 150 200 250
Number at risk
IA−S1 325 159 19 2 0 0
IA−S2 301 101 9 0 0 0
IB−S1 240 102 14 2 1 0
IB−S2 314 116 13 0 0 0
0 50 100 150 200 250
Time (Month)
D
Relapse-free Survival
1.00 0.75 0.50 0.25 0.00
HR = 2.91
P < 0.0001
0 30 60 90 120
Number at risk
IA−S1 113 101 55 8 0
IA−S2 92 71 34 3 0
IB−S1 48 36 24 3 0
IB−S2 88 48 28 6 0
0 30 60 90 120
Time (Month)
IA−S1
IA−S2
IB−S1
IB−S2

**FigureS5**

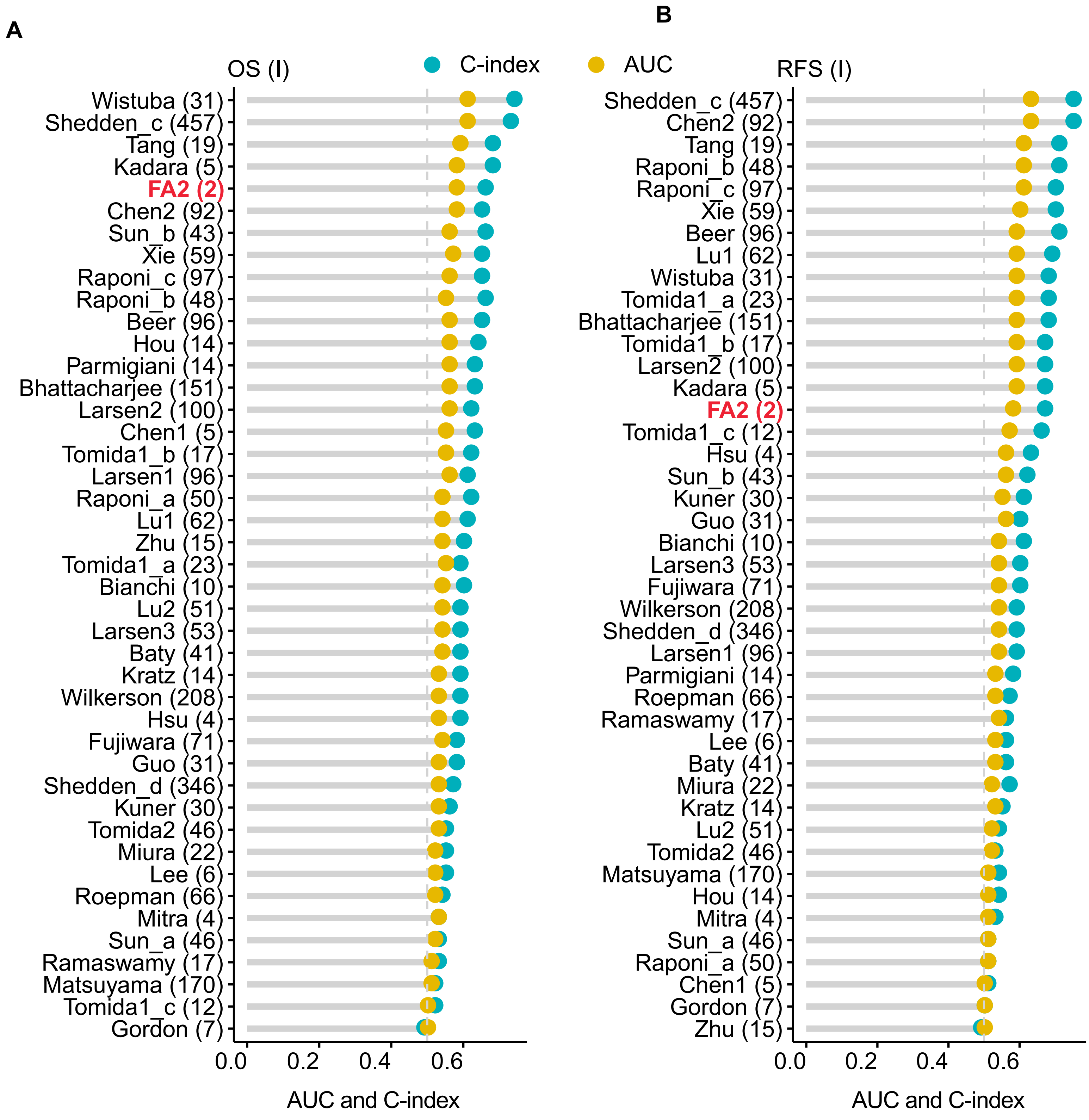
A
B
OS (I)
C-index
AUC
RFS (I)
Wistuba (31)
Shedden_c (457)
Tang (19)
Kadara (5)
FA2 (2)
Chen2 (92)
Sun_b (43)
Xie (59)
Raponi_c (97)
Raponi_b (48)
Beer (96)
Hou (14)
Parmigiani (14)
Bhattacharjee (151)
Larsen2 (100)
Chen1 (5)
Tomida1_b (17)
Larsen1 (96)
Raponi_a (50)
Lu1 (62)
Zhu (15)
Tomida1_a (23)
Bianchi (10)
Lu2 (51)
Larsen3 (53)
Baty (41)
Kratz (14)
Wilkerson (208)
Hsu (4)
Fujiwara (71)
Guo (31)
Shedden_d (346)
Kuner (30)
Tomida2 (46)
Miura (22)
Lee (6)
Roepman (66)
Mitra (4)
Sun_a (46)
Ramaswamy (17)
Matsuyama (170)
Tomida1_c (12)
Gordon (7)
0.0
0.2
0.4
0.6
AUC and C-index
Shedden_c (457)
Chen2 (92)
Tang (19)
Raponi_b (48)
Raponi_c (97)
Xie (59)
Beer (96)
Lu1 (62)
Wistuba (31)
Tomida1_a (23)
Bhattacharjee (151)
Tomida1_b (17)
Larsen2 (100)
Kadara (5)
FA2 (2)
Tomida1_c (12)
Hsu (4)
Sun_b (43)
Kuner (30)
Guo (31)
Bianchi (10)
Larsen3 (53)
Fujiwara (71)
Wilkerson (208)
Shedden_d (346)
Larsen1 (96)
Parmigiani (14)
Roepman (66)
Ramaswamy (17)
Lee (6)
Baty (41)
Miura (22)
Kratz (14)
Lu2 (51)
Tomida2 (46)
Matsuyama (170)
Hou (14)
Mitra (4)
Sun_a (46)
Raponi_a (50)
Chen1 (5)
Gordon (7)
Zhu (15)
0.0
0.2
0.4
0.6
AUC and C-index

**FigureS6**

Table S1: Summary of data sets used in this study

| No | Data set ID | Omics data type | Platform | No. of stage I LUAD patients | Patient ratio (S1/S2) | References |
|---|---|---|---|---|---|---|
| **1** | EGAS0000 1004006 | Transcriptomic | Illumina HiSeq X | 83 | 1.52 | 1 |
| **2** | GSE31210 | Transcriptomic | Affymetrix Human Genome U133 Plus 2.0 Array | 168 | 0.89 | 2,3 |
| **3** | GSE13213 | Transcriptomic | Agilent-014850 Whole Human Genome Microarray 4x44K G4112F | 79 | 1.32 | 4 |
| **4** | GSE11969 | Transcriptomic | Agilent Homo sapiens 21.6K custom array | 47 | 1.94 | 5,6 |
| **5** | GSE50081 | Transcriptomic | Affymetrix Human Genome U133 Plus 2.0 Array | 92 | 0.56 | 7 |
| **6** | GSE68465 | Transcriptomic | Affymetrix Human Genome U133A Array | 108 | 1.16 | 8 |
| **7** | GSE42127 | Transcriptomic | Illumina HumanWG-6 v3.0 expression beadchip | 89 | 0.68 | 9,10 |
| **8** | GSE26939 | Transcriptomic | Agilent-UNC-custom-4X44K | 62 | 1.48 | 11 |
| **9** | GSE72094 | Transcriptomic | Rosetta/Merck Human RSTA Custom Affymetrix 2.0 microarray | 265 | 0.74 | 12 |
| **10** | GSE30219 | Transcriptomic | Affymetrix Human Genome U133 Plus 2.0 Array | 71 | 0.54 | 13 |
| **11** | GSE41271 | Transcriptomic | Illumina HumanWG-6 v3.0 expression beadchip | 100 | 0.67 | 14-17 |
| **12** | TCGA | Genomic | Illumina HiSeq 2000 | 232 | - | 18 |
| **12** | TCGA | Transcriptomic | Illumina HiSeq 2000 | 232 | 1.13 | 18 |
| **13** | Gillette et al | Genomic | Illumina HiSeq4000/ HiSeqX | 55 | - | 19 |
| **13** | Gillette et al | Transcriptomic | Illumina HiSeq4000 | 55 | 0.28 | 19 |
| **13** | Gillette et al | Proteomics | LC-MS/MS | 55 | - | 19 |
| **13** | Gillette et al | Phosphorylated proteomics | LC-MS/MS | 55 | - | 19 |
| **14** | Xu et al | proteomics | LC-MS/MS | 51 | - | 20 |

Table S2: Summary of clinical features of datasets of stage I lung adenocarcinoma patients without adjuvant treatment

| **Clinical** | **FUSCC** | **GSE31210** | **GSE13213** | **GSE11969** | **GSE50081** | **GSE68465** | **GSE42127** | **GSE26939** | **GSE72094** | **GSE30219** | **GSE41271** | **TCGA** | **Gillette** | **Xu** |
|---|---|---|---|---|---|---|---|---|---|---|---|---|---|---|
| **No. Patients** | 83 | 168 | 79 | 47 | 92 | 108 | 89 | 62 | 265 | 71 | 100 | 232 | 55 | 51 |
| **Gender** | | | | | | | | | | | | | | |
| Male (%) | 39 (47.0) | 71 (42.3) | 41 (51.9) | 27 (57.4) | 46 (50.0) | 44 (40.7) | 38 (42.7) | 36 (58.1) | 122 (46.0) | 55 (77.5) | 44 (44.0) | 100 (43.1) | 35 (63.6) | 33 (64.7) |
| Female (%) | 44 (53.0) | 97 (57.7) | 38 (48.1) | 20 (42.6) | 46 (50.0) | 64 (59.3) | 51 (57.3) | 26 (41.9) | 143 (54.0) | 16 (22.5) | 56 (56.0) | 132 (56.9) | 20 (36.4) | 18 (35.3) |
| **Age** | | | | | | | | | | | | | | |
| < 60 (%) | 34 (41.0) | 71 (42.3) | 26 (32.9) | 13 (27.7) | 13 (14.1) | 30 (27.8) | 32 (36.0) | 18 (29.0) | 36 (13.6) | 33 (46.5) | 37 (37.0) | 62 (26.7) | 21 (38.2) | 22 (43.1) |
| >= 60 (%) | 49 (59.0) | 97 (57.7) | 53 (67.1) | 34 (72.3) | 79 (85.9) | 78 (72.2) | 57 (64.0) | 44 (71.0) | 229 (86.4) | 38 (53.5) | 62 (62.0) | 156 (67.3) | 34 (61.8) | 29 (56.9) |
| Unknown | | - | - | - | - | - | - | - | - | - | 1 (1.0) | 14 (6.0) | - | - |
| **Smoking** | | | | | | | | | | | | | | |
| Never (%) | 59 (71.1) | 94 (56.0) | 39 (49.4) | 24 (51.1) | 20 (21.7) | - | - | 7 (11.3) | 20 (7.5) | - | 16 (16.0) | 30 (12.9) | - | - |
| Ever (%) | 24 (28.9) | 74 (44.0) | 40 (50.6) | 23 (48.9) | 63 (68.5) | - | - | 55 (88.7) | 208 (78.5) | - | 84 (84.0) | 201 (86.7) | - | - |
| Unknown | | - | - | - | 9 (9.8) | - | - | - | 37 (14.0) | - | - | 1 (0.4) | - | - |
| **Stage** | | | | | | | | | | | | | | |
| IA (%) | 56 (67.5) | 114 (67.9) | 42 (53.2) | 25 (53.2) | 36 (39.1) | - | 32 (36.0) | 31 (50.0) | 158 (59.6) | - | 36 (36.0) | 110 (47.4) | 21 (38.2) | 16 (31.4) |
| IB (%) | 27 (32.5) | 54 (32.1) | 37 (46.8) | 22 (46.8) | 56 (60.9) | - | 57 (64.0) | 31 (50.0) | 101 (38.1) | - | 64 (64.0) | 118 (50.9) | 32 (58.2) | 35 (68.6) |
| I (%) | - | - | - | - | - | 108 (100.0) | - | - | 67 (2.3) | 71 (100.0) | - | 4 (1.7) | 2 (3.6) | - |
| **Subtype (FA2)** | | | | | | | | | | | | | | |
| S1 (%) | 50 (60.2) | 79 (47.0) | 45 (57.0) | 31 (66.0) | 33 (35.9) | 58 (53.7) | 36 (40.4) | 37 (59.7) | 113 (42.6) | 25 (35.2) | 40 (40.0) | 123 (53.0) | 12 (21.8) | - |
| S2 (%) | 33 (39.8) | 89 (53.0) | 34 (43.0) | 16 (34.0) | 59 (64.1) | 50 (46.3) | 53 (59.6) | 25 (40.3) | 152 (57.4) | 46 (64.8) | 60 (60.0) | 109 (47.0) | 43 (78.2) | - |
| **RFS** | | | | | | | | | | | | | | |
| Yes (%) | 36 (43.4) | 37 (22.0) | 32 (40.5) | 15 (31.9) | 22 (23.9) | 31 (28.7) | - | - | - | 17 (23.9) | 30 (30.0) | - | 5 (9.1) | 30 (58.8) |
| No (%) | 47 (56.7) | 131 (78.0) | 46 (58.2) | 31 (66.0) | 68 (73.9) | 53 (49.1) | - | - | - | 54 (76.1) | 70 (70.0) | - | 37 (67.3) | 21 (41.2) |
| Unknown | - | - | 1 (1.3 %) | 1 (2.1) | 2 (2.2) | 24 (22.2) | - | - | - | - | - | - | 13 (23.6) | - |
| **OS** | | | | | | | | | | | | | | |
| Alive (%) | 74 (89.2) | 151 (89.9) | 54 (68.4) | 34 (72.3) | 61 (66.3) | 74 (68.5) | 67 (75.3) | 31 (50.0) | 208 (78.5) | 38 (53.5) | 74 (74.0) | 182(78.4) | 51 (92.7) | 43 (84.3) |
| Dead (%) | 9 (10.8) | 17 (10.1) | 25 (31.6) | 13 (27.7) | 31 (33.7) | 34 (31.5) | 22 (24.7) | 31 (50.0) | 57 (21.5) | 33 (46.5) | 26 (26.0) | 45 (19.4) | 4 (7.3) | 8 (15.7) |

RFS: relapse-free survival
OS: overall survival

Table S3: Gene lists of 43 prognostic predictive models

| Models (Number of Genes) | List of Genes |
|---|---|
| Baty (41) | ARPC2, FOXQ1, SDF2, MOSPD3, PMS2L9, MUT, NOC4, NDUFC1, KIAA2010, AP3D1, XP_372900.2, MRPL44, CHCHD2, SLC37A2, ACTR10, Q6PIE2, TCEB3, INO80, MYO1E, LOC90410, APG5L, MGC33302, FTSJ1, VEGFB, RNF103, CYB561D2, OPTN, MAT2B, HEBP2, LRRC9, CSNK1A1, CLIP1, MUS81, ARG2, SNAP29, LOC91661, NYREN18, MGC10067, RMI1, MBTPS1, CBWD2 |
| Beer (96) | BAG1, CASP4, FADD, P63, 5T4, ITGA2, KRT18, KRT19, KRT7, LAMB1, TMSB4X, TUBA1, BMP2, CDC6, H2AFZ, PDAP1, POLD3, REG1A, S100P, SERPINE1, STX1A, ADM, AKAP12, ARHE, DEFB1, GRB7, INHA, ITK, NACA, STC1, TNFAIP6, VEGF, VLDLR, WNT1, WNT10B, HSPA8, ERBB2, FXYD3, HLA-B, HPCAL1, P2RX5, PEX7, SLC20A1, SLC2A1, VDAC2, ALDH8, ALDOA, ATP2B1, CDS1, CSTB, CTSL, CYP24, FUCA1, FUT3, GAPD, GCNT1, HMBS, KYNU, MLN64, MSH3, MT2A, NME2, NP, PACE, PDE7A, PLGL, PPIF, PTPRCAP, RPC, SC4MOL, SLC1A6, UBC, UGP2, UQCRC2, COPEB, CRK, DBP, GARS, HRB, PRDM2, RELA, RPS26, RPS3, RPS6KB1, SUI1, TIEG, TMF1, FEZ2, HPIP, KIAA0005, KIAA0020, KIAA0084, KIAA0153, KIAA0263, KIAA0317, MGB1 |
| Bhattacharjee (151) | CDX2, CDH17, CYP3A5, CDX1, PRSS3, HEPH, NOX1, FABP1, PLS1, CEACAM1, MTMR11, ATP10B, SATB2, LGALS4, KRT20, HSD11B2, CLDN3, PTK6, ITGA6, VIL1, ETHE1, USH1C, MYO1A, GMPS, MCM6, SNRPG, BUB3, RFC4, PFN2, HMGB2, TUBB, FEN1, PCNA, H2AFX, TPX2, STIP1, SMC3, TTK, UBE2S, CDC2, TCP1, CKS2, UNG, KLK11, ASCL1, CPE, TFF3, CALCA, PCSK1, HSP90B1, TUBB2B, XBP1, GRAMD4, MED13L, KIAA1128, NFYB, DUSP4, ZWINT, FOXA1, DDC, PAFAH1B3, RET, TOX3, ATP1B1, AHCYL2, PGC, ODC1, GSTP1, S100P, SFTPB, ALDH3A2, AKR1C1, AKR1C2, PLA2G10, SCNN1A, PDE4D, NQO1, EVI1, SMPDL3B, CBR1, MUC1, PLA2G4A, PDIA3, ALDH3B1, TCF25, PITRM1, FHL1, TGFBR2, CAV1, AGER, COX7A1, PECAM1, DPYSL2, TACC1, MARCO, ICAM2, CLEC3B, MFAP4, CFD, PTRF, AHNAK, HEG1, AKAP2, RAMP2, SERPING1, GPX3, QKI, AOC3, STOM, CYP11B1, CTSH, FOLR1, GPD1L, PTPN4, MALL, FRMD4B, MTRR, CD74, HSD17B6, PTPN13, CYB5A, SSTR4, ANK3, NKX2-1, ROS1, ACSM3, THUMPD1, MGLL, HLA-DOA, CDKL2, NR3C2, CSNK1D, NPEPPS, VEGFA, MYOF, AMIGO2, ITGA3, ABCC3, PHLDA2, SDC1, DGCR2, CDC42EP1, CCND1, SMURF2, CDC42BPA, ADK, APLP2, CHST3, PON2, PTK2, TOM1L1 |
| Bianchi (10) | NUDCD1, E2F1, HOXB7, MCM6, SERPINB5, E2F4, HSPG2, SF3B1, RRM2, SCGB3A1 |
| Chen1 (5) | DUSP6, MMD, STAT1, ERBB3, LCK |
| Chen2 (92) | AMOTL2, APOBEC3B, ARF1, ARF4, ARFGEF1, ASPM, BIRC5, BUB1, BUB1B, BUB3, C10orf3, CBX7, CCNB1, CCNB2, CCNE2, CDC2, CDC20, CDKN1C, CDKN3, CENPA, CENPF, CHRDL1, CKAP2, CKS2, CORO2A, CTPS, DKFZp762E1312, DLG7, DONSON, DPP3, DST, DTL, DXS9879E, ECM2, ECT2, EZH2, FLJ10719, FOXM1, GPC3, GPRASP1, H2AFX, H2AFZ, HIST1H2BF, HMGB3, HMMR, HN1, KIAA0101, KIF11, KIF20A, KIF23, KIF4A, KNTC2, KPNA2, LEPR, MAGI2, MAPK13, MCM2, MCM4, MELK, MLF1IP, MOBK1B, MRPL42, MT1M, NCBP1, NEK2, NME1, NUP210, NUSAP1, PAFAH1B3, PBK, PCNA, PDZK3, PPIF, PRC1, PSF1, RACGAP1, RAD51AP1, RRM2, SMC2L1, SMC4L1, SPP1, SQLE, STK6, TIMELESS, TK1, TOP2A, TPX2, TTK, UBE2C, UBE2S, ZWILCH, ZWINT |
| FA2 (2) | COL11A1, THBS2 |
| Fujiwara (71) | ASCL1, TCERG1L, NOL4, DPYSL5, KIAA1456, PCP4, XIST, RGS4, PTER, CDKN2C, PSIP1, CDKN2D, FOXN4, GRP, NP25, SCGB2A1, FAM33A, MMP12, MAGI1, EVC, CP110, CNTNAP2, SEC11L3, HOXC5, SEZ6L, DPP10, ELAVL4, ANKRD32, CHD7, OXCT2, PPM1E, WHSC1, SEZ6, HMGN2, CAMK2B, LPAAT-e, CDCA1, FOXG1B, MYCL1, KIAA1853, FARP1, LHX2, DDX11, MAGEB3, MSI1, SLC36A4, SKP1A, TSGA14, ZNF385B, CA8, M11S1, TOP2A, GHRH, MED25, MGC57827, HOXB5, FRAS1, CENPF, ATXN7L4, STMN1, TUBA3, ARID5B, ID4, CRLF3, CDK5R1, ENO2, NAV2, MYEF2, GTSE1, ELMOD2, RAB3B |

| | |
|---|---|
| Gordon (7) | TRB@, APOE, LPIN2, SLC2A1, S100P, MST1R, FXYD3 |
| Guo (31) | CREB3, RER1, FCN2, NP220, FBRNP, LBC, MSX2, TAL2, GHRHR, MT3, TNFSF9, ARHGDIG, TUBA3, EGF, HFL3, FUT7, GUCA2B, ATP5A1, EZFIT, UBE1, ATRX, ILF3, E2F4, TAX1BP2, UBE2I, OGT, INSR, GNB1, CHD4, EMK1, HRMT1L2 |
| Hou (14) | DGKH, EIF5A, CPT1A, ADH1C, BNC1, HSD17B3, XPO7, ENPP2, COX8C, EXOSC6, EGFR, RNASE7, C12orf59, MYNN |
| Hsu (4) | ANKRD49, LPHN1, RABAC1, EGLN2 |
| Kadara (5) | UBE2C, MCM2, MCM6, FEN1, TPX2 |
| Kratz (14) | BAG1, BRCA1, CDC6, CDK2AP1, ERBB3, FUT3, IL11, LCK, RND3, SH3BGR, WNT3A, ESD, TBP, YAP1 |
| Kuner (30) | DSG3, S100A7, GJB5, GJB6, PVRL1, CLDN1, DSC2, CLDN3, CLDN7, CLDN12, CLDN23, PARD6B, TJP3, CGN, CDC42BPA, PARD6G, DLG1, MUC1, GSC, SNAI2, TWIST1, MMP9, CDH1, CDH2, ZEB2, SNAI1, ZEB1, TWIST2, POLR2A, ESD |
| Larsen1 (96) | PDCD10, BITE, RHBDF1, RAB11FIP4, SCGB2A1, HRASLS, FLJ21836, DCLRE1C, PTPRC, EVPL, CDH20, TPBG, PDZD2, DOCK2, C11orf10, CRISP3, ITCH, HSPG2, LMLN, TMEM47, KLHL3, DYM, FRMD4A, SCGB1D1, LRIG2, C3orf9, C18orf15, STX11, STEAP1, GOLPH3L, PP35, P38IP, FAM11A, TRSPAP1, SFRS10, PTPLA, IL7R, KCNMA1, SEL1L, ASB7, PRLR, ARFGEF2, TIAM1, CABP3, COX15, ABCA1, PTPN21, HCDI, RDH14, PPM1B, KIAA0372, ENPP6, ACOT2, CLPX, CAPN3, RSPRY1, ZNF292, TRIM22, ZNF277, POLR3G, NR1D2, CHD2, IMP4, ATP6N1A, C1QC, FTL, NP, SLC37A1, FXYD3, TRPM1, ABCC5, AMDHD1, RPL15, TRPC5, CES1, EML2, MAF, BRWD1, CTSS, HERC2P7, C1orf112, MGC16037, FLJ31943, C16orf57, LOC129285, C1orf86, RP11-308B5.5, PGBD5, CSMD2, PIN1L, LOC144438, C14orf155, PRO1051, ZNF302, LOC92482, LOC642776 |
| Larsen2 (100) | DNAJB12, MGC20255, MGC4767, P2Y5, ACCN3, FLNC, NAG-7, SGCD, KNSL4, ERP70, PDE3A, ZFP106, FLJ13373, FLJ10430, PABPC5, CD83, LRP2, VARS2, LOC51042, CDX4, GFRA2, OPRM1, PTPRE, NR1D2, ZNF197, FLJ11011, CHST2, CYP3A7, CRP, DUSP9, RDBP, KRTHB1, FLJ12606, SLC2A10, PVRL2, SPRY4, ENTPD6, AD036, FLJ11068, GPX5, MKLN1, TGFBR1, ZNF228, HAP1, PLAB, PODLX2, DELGEF, FLJ23529, C5orf5, FLJ23185, BIRC3, DKFZP434I2117, FLJ10773, C20orf72, BASP1, GRHPR, KIAA1373, ATP6L, AIP, ARF3, FLJ21657, LOC51114, SSRP1, FLJ11045, MGC15737, FLJ12610, IGLJ3, KIAA0707, MGC3260, SPS, SEPT6, ETS1, GTR2, FLJ13381, PRO1693, CLUL1, ZNF-U69274, COX6BP-3, FLJ23093, SNX17, IFNA2, NTN4, SYTL2, ITGAX, RGS5, STK18, IGLV9-49, KIAA0138, MRPL3, KIAA1718, KIAA1920, MGC2601, ATP8B2, GBX2, H3FK, ADAMTS1, MMD, TBX19, EPO, KIAA0644 |
| Larsen3 (53) | IL8, CLU, API5, OPA1, GULP1, ARMCX3, ANXA4, ZPBP, KRTHB4, FHOD3, POLG2, FBXO32, CDC23, MGP, HGFAC, PKP3, JPH4, PCDHGC3, AKAP4, RSBN1, ODF2, CYB5A, CPNE1, SIAE, FLJ20628, DDOST, PTPRH, RPS4Y1, BRAP, EBI2, DUSP5, LOC375449, MARK3, ZNF8, ZBTB1, NFIX, PRNP, SMCY, ZNF268, ZNF198, ZBTB11, ATXN3L, LOC152485, MGC21675, TMEM155, RHBDD1, FLJ20054, LOC648232, VIT, LOC644710, RACGAP1P, FLJ11734, PRO2964 |
| Lee (6) | CALB1, MMP7, SLC1A7, GSTA1, CCL19, IFI44 |
| Lu1 (62) | APC, ARHGEF1, BCL2, CASP10, CASP8, CDH8, DTNA, ENPP2, GOLGA1, IL8RB, ITSN1, NTRK3, PPOX, RAB28, RAE1, SON, CCR2, FUCA1, GNAT2, MAP4K1, MEF2C, MLLT10, NR1H4, PBP, PIGC, PIK3R1, PKNOX1, PRKACA, SNX1, TMSB4X, TRA2A, ZNFN1A1, ABCC1, ADAM17, BLM, CHERP, CRABP1, INHA, LY6D, NID, NOTCH3, PFN2, PSEN1, SMC1L1, STC1, ARL4A, BIK, DSP, FBN2, GLI2, HNRPD, IRS1, LARS2, PLEC1, PYGL, SLC2A1, SLC7A1, SMARCA3, UPK2, VGLL1, ZNF154, ZNF410 |
| Lu2 (51) | AU148154, B4GALT1, CGB, CHST12, CLEC11A, COL2A1, CYP2A6, DENND1A, DIO1, DOCK6, EPHB6, FZD9, GLE1, GTF3C2, INF2, KDM4B, SIK3, GREB1L, KLK5, KRT81, LENEP, MYOG, NFKBIL1, NLRP2, NM_004876, FEZ2, NM_018600, OCA2, PADI3, RPRM, SH3YL1, SLC27A2, SLC35F5, SNAPC2, SPTBN2, STRN3, SUSD4, TCF3, TET3, THBS1, TRIM34, TRIM46, TRIP11, CELSR1, UBE2D4, UBXN4, VKORC1, ZBTB7B, ZNF365, MUC5AC, FGFR2 |

| | |
|---|---|
| Matsuyama (170) | SFTPB, PLCL1, KLK4, CAMK4, PHACTR1, THAP4, NPFF, CTLA4, KCNQ1DN, GRID2, FABP4, SLC34A2, PCAF, SEMA4G, BGR, NIP, ABCA3, RAB33B, HAS3, SFTPG, ZNFN1A1, ATP10A, CGI-38, SCML2, STK32B, OR51E2, FLJ12476, FAM69B, APOL5, SAP30L, DEPDC2, COL20A1, SERPINB10, SPOCD1, FAM76A, CLU, CAMKK1, GPR92, ATRX, THAP7, FGF18, LOC162427, CRISPLD2, PLXNB1, KIAA1279, E2F3, FLJ12057, MLL2, HEMGN, TNFAIP8L1, ZNF613, KCNJ15, ZBTB12, ATP10B, SLC5A5, TMEM37, STXBP2, RAD9A, TRIM54, AGBL4, ITGA2B, TEX13B, TCTA, CPLX1, ETV2, IQCE, LIP8, PGK1, HSD11B2, HLA-DRB1, KIAA0420, C15orf16, SNCG, IMPDH2, PCDH21, FYN, C1orf71, IRAK2, KIAA1456, FUT8, ZHX2, PDP2, FLJ37300, NDST1, N4BP1, DOM3Z, JAK2, CRLF2, OR52E5, CA13, CYP2A13, RAB11FIP2, BARHL2, ZG16, ACP2, XPO6, MAFK, PNCK, CACNA1I, LRRC14, HIVEP3, XYLB, DRD2, ADCY5, MATK, LMO2, CDGAP, SLC7A6, DEFB129, CFHL2, CYP17A1, FBXL14, PRAMEF8, HIP1, LRRC7, FUT10, DLX6, SKI, UBXD5, MIOX, CD300LB, VTI1A, CST1, KIAA1409, SCTR, DHDDS, FLJ40083, NAP1L4, C10orf87, SCGB1A1, SERPINA1, AVPR1B, ANKRD41, ESR1, MGC13057, NGFB, PDE4D, MLN, C21orf25, PSG9, MMRN2, MGC21644, CDK8, TSPY1, USH3A, CMYA4, PLA2G2F, SFTPD, LOXHD1, UNQ3045, HRMT1L3, CYP4B1, GOLGA6, ING1, RRAD, KIF5A, DMBT1, PIK3AP1, SLC22A16, LEMD1, APOE, CDKL2, PCDH11Y, GGTLA4, MMP28, KLK3, OLFM1, AQP5, COL10A1, HOP |
| Mitra (4) | DBN1, FLAD1, CACNB3, CCND2 |
| Miura (22) | BUB3, ZW10, DCTD, KRT9, PSMC5, EYA2, NXF1, AUP1, NDUFS3, PSMC1, ANAPC2, ASMTL, RPL24, COX14, TESPA1, A1BG, ASB6, TPRA1, ESYT1, NOP2, NCKIPSD, SMAP2 |
| Parmigiani (14) | GPC3, MALL, IRX5, FGFR2, FOLR1, TYRP1, STX1A, IGJ, MAD2L1, VEGFC, KIAA0101, IL6ST, SELE, ARHGDIB |
| Ramaswamy (17) | SNRPF, EIF4EL3, HNRPAB, DHPS, PTTG1, COL1A1, COL1A2, LMNB1, ACTG2, MYLK, MYH11, CNN1, HLA-DPB1, RUNX1, MT3, NR4A1, RBM5 |
| Raponi_a (50) | ADM, AKAP12, ALDH3B2, BZW1, CASP4, CDC6, CKAP4, CRK, CSTB, EIF1, FADD, FEZ2, FURIN, FUT3, FXYD3, GAPDH, GARS, GRB7, H2AFZ, HPCAL1, IHPK1, INHA, KIAA0020, KLF6, KRT18, KRT7, MT2A, NP, PEX7, RAFTLIN, RND3, RPS3, RTCD1, S100P, SCGB2A2, SCL20A1, SCL2A1, STARD3, STC1, TMSB4X, TUBA1, UGP2, UQCRC2, WNT10B, ERBB2, HRB, TPBG, ATP2B1, PLGLB2, TMF1 |
| Raponi_b (48) | TIA1, BID, MAP4, CASK, EVI1, PPFIBP2, MS4A1, RAB6A, RAB6C, AGFG1, PTPN18, CCL11, FGFR2, MAPK14, MARK1, GGA3, LPAR1, NPHP4, RAB11A, LGALS8, AASS, HMBS, ABCC4, DPAGT1, FAM108B1, PPME1, ALG8, PSMC2, CPA3, ANKRD12, SPAST, ZNF552, ZNF189, RPLP0, BMS1, HLF, NARS2, TMEM126B, LRIG1, PELI2, TAF1D, IGL@, HEATR2, YPEL5, C1orf115, KIAA0746, SERINC2, HRB |
| Raponi_c (97) | ADM, AKAP12, ALDH3B2, BZW1, CASP4, CDC6, CKAP4, CRK, CSTB, EIF1, FADD, FEZ2, FURIN, FUT3, FXYD3, GAPDH, GARS, GRB7, H2AFZ, HPCAL1, IHPK1, INHA, KIAA0020, KLF6, KRT18, KRT7, MT2A, NP, PEX7, RAFTLIN, RND3, RPS3, RTCD1, S100P, SCGB2A2, SCL20A1, SCL2A1, STARD3, STC1, TMSB4X, TUBA1, UGP2, UQCRC2, WNT10B, ERBB2, HRB, TPBG, ATP2B1, PLGLB2, TMF1, TIA1, BID, MAP4, CASK, EVI1, PPFIBP2, MS4A1, RAB6A, RAB6C, AGFG1, PTPN18, CCL11, FGFR2, MAPK14, MARK1, GGA3, LPAR1, NPHP4, RAB11A, LGALS8, AASS, HMBS, ABCC4, DPAGT1, FAM108B1, PPME1, ALG8, PSMC2, CPA3, ANKRD12, SPAST, ZNF552, ZNF189, RPLP0, BMS1, HLF, NARS2, TMEM126B, LRIG1, PELI2, TAF1D, IGL@, HEATR2, YPEL5, C1orf115, KIAA0746, SERINC2 |
| Roepman (66) | C3orf41, C1orf24, PLEK, RHOH, PSCDBP, TCEA2, FLJ21963, CLEC4E, USP51, WFDC10B, IGH@, SLC4A3, CD53, THRAP2, PRDM13, OBSL1, TAGAP, MGC11271, IGLV6-57, CD38, FKBP9, ADAMTSL2, CD48, GNPTAB, DHRSB, LOC388886, CNIH3, PSMA6, CCRK, SHROOM1, GPSM1, TRO, GSTT2, NQ02, EAF2, MUM1L1, MUC4, C13orf21, PABPC1, PLA2G7, PARK2, AOAH, IGL@, TMSB4X, ACOT8, GIMAP7, ASAH1, TRIM45, C2orf30, EXT2, IFI6, KCNE3, CTSF, SULT1C1, RASL11B, LOC148898, HMGCL, IGHA1, C1QTNF3, C20orf46, IL11RA, ADRA2C, IGKC, CEACAM5, PURB, TPD52 |

| | |
|---|---|
| Shedden_c (457) | LDHC, HCCS, LDHB, XRCC3, SLC9A2, SYNCRIP, ANKLE2, CDCA8, WDR76, ZNF248, HIST1H2BJ, PHTF2, YEATS2, LSM4, CCNA2, CDCA3, KRR1, TMEFF1, TFPT, C16orf61, MTPAP, NCAPD3, ORC1L, NCAPD2, NME1, SPAG5, ZWINT, FOXG1, YME1L1, KIF4A, GNAI3, CCDC93, ZSCAN5A, UBA6, ESM1, MYBL1, MTIF2, ARFGEF2, MYBL2, ACP1, MAGEA6, PFN2, HEXIM1, NCAPG2, NAT13, C19orf28, RECQL4, GPD2, YEATS4, MKI67, GGH, NDC80, WDR67, NOLC1, WDR62, PKP2, PCNA, PPP2R3C, SEPHS1, AP2S1, EZH2, GTSE1, CCNE2, CCNE1, MTHFD2, IL1RAP, ANGPT2, CDC7, CDC6, CDC2, TTC38, TPX2, EXOSC2, HN1, DGUOK, TTF2, MMP12, CIAO1, EIF4A3, SS18, RRM2, NAB1, GNAS, RUVBL2, ARL4D, WDR43, HIST1H2AG, HK2, AK3L1, CDC45L, NUP210, BUB1, RQCD1, BUB3, TRIP13, HIST1H2BH, PHF10, CPT1A, CCNB1, MPHOSPH9, NRAS, CCNB2, CSNK1D, RAB22A, DUSP9, PSAT1, YKT6, NCBP1, IMPAD1, RNMT, PRC1, DBF4, HBS1L, KNTC1, SLC7A5, EIF4EBP1, CDKN2A, ATG5, OIP5, CDKN2C, ELOVL4, DNAJC9, CDKN2D, SLC2A1, RALA, TUBG1, ELOVL6, SAR1A, ASPM, SNRPA1, TWF1, NUDT3, GOLT1B, CMAS, NEIL3, RBL1, TACC3, PA2G4, STMN1, LRPPRC, FBXO11, NETO2, HMGB2, MAPKAPK5, UBE2V2, CCL7, QSER1, POLE2, DPY30, MEMO1, TEAD4, DNAJA1, HELLS, TFDP1, ERCC6L, GINS1, GINS2, GINS3, GINS4, ARPP19, WAPAL, BRIP1, WHSC1, MOBKL1B, R3HDM1, DNA2, PLK4, HDAC2, GPR37, KIF23, NBN, E2F8, CAD, PAWR, KIF2C, WARS, ACOT7, PRMT1, PAX8, MAGOHB, DDA1, KIF2A, KIF14, SEC23A, KIF11, IL18RAP, GPR19, KIF15, NUSAP1, PBK, HMGA1, TMEM38B, PPM1G, RECQL, RIF1, IPO5, ERN1, HAUS3, TMEM194A, FKBP4, EGLN3, DCK, WDYHV1, GPSM2, EXO1, C13orf27, SMC6, CDC25C, SMC2, XPNPEP1, CDC25A, SLBP, CBS, PVR, SNCG, C7orf44, MRPL42, THOP1, PTTG2, AURKA, AURKB, PTTG1, TAF7L, ATP2B1, PRIM1, PLOD2, PRIM2, H2AFZ, CHRNA5, NUP37, H2AFX, PITX1, RPP25, F12, STC2, VANGL1, RAN, STRN4, C1orf103, TBRG4, DEPDC1, PNKP, PGM3, RFC3, MAD2L1, RFC4, MTF2, RFC2, VEGFA, TUBA4A, STC1, CTSL2, CDK5R1, FIP1L1, LMNB1, ENPP1, NEK2, C6orf120, CHEK1, CD70, SFXN1, IGF2BP3, PPAT, TK1, ACD, VRK2, FBXO5, SKA1, WDHD1, FCHO1, RTCD1, SCD, DENND1A, CDC20, RAD54L, CDC27, OBFC2B, FAM64A, CCT5, NUP62, PPARD, BOLA2, LOC440354, MYO7A, WASF1, CDT1, ACTR3, AP2B1, AGPAT5, C12orf24, FANCI, SEMA3A, TOP2A, FANCA, C1orf144, DSN1, ARTN, ADM, ROD1, MED8, CARHSP1, FUT9, PPFIA1, TFG, COL2A1, HMMR, GCH1, MIF, NCAPH, C12orf48, MRPL13, MRPL12, PLIN2, KLC1, NCAPG, MRPL19, PCMT1, PRKAA2, UGT8, ZWILCH, TPRKB, NMU, NA, POLR3G, DLGAP5, BIRC5, CDKN3, SOD2, , SLC16A3, AFP, PSMD12, PYGL, CALM3, PGK1, PAICS, KIF20A, CLPB, DTYMK, EIF5B, TTK, PMAIP1, MCM10, TSSC1, SMNDC1, CBX5, APOBEC3B, GABPB1, SLC16A1, RC3H2, ITCH, DARS, DTL, TIMM8A, ADRM1, TNNT1, PSMA5, FGFR1OP, SNRPB, RIPK2, DSP, SLC38A1, MAP6D1, PRPS1, TWSG1, SNX6, SRM, SNX7, UBAC1, PSMA7, COMMD8, PRR7, RASAL2, SPC25, CSE1L, MTCH2, C12orf11, PSMB2, ERO1L, SSX2IP, UBE2D1, LRFN4, CKAP2, TSR1, TOMM40, ATAD2, BRCA2, KCTD5, PPIF, KLRC1 , KLRC2, SLC25A10, PPID, TROAP, GLMN, CORT, EIF4E2, PDCD2, GSS, SAP30, POMT2, SRD5A1, PFKP, UBE2C, MCM4, MCM5, MCM6, C20orf20, UBE2N, UBE2K, C2orf3, BUB1B, PSME3, KPNA2, UBE2S, MELK, SHCBP1, FOXM1, KIAA0101, PRKDC, HAT1, CEP55, CCDC59, TYMS, CDYL, C11orf9, CENPA, HJURP, TUBA3C, TUBA3D, GAPDH, KRT8, LOC149501, MSH6, CENPN, CENPM, TCP1, DNM1L, RAD51AP1, IL2RA, NDUFA9, CENPQ, PSRC1, KIF18A, KIF18B, CENPF, CENPE, RACGAP1, MLF1IP, CENPI, GMFB, RGS20, KAZALD1, DR1, CKS2, STEAP1B, SETD8, MUC16 |
| Shedden_d (346) | ADORA3, GPR126, FAM20B, SAA3P, L1CAM, VPRBP, C1orf113, AQP6, IGHM, C14orf56, BBOX1, ISG20, FAM30A, VPS13D, KIAA0125, IGLV6-57, RAB26, IGL@, IGLV1-44, MAP2K7, IGLL3, SH3GL3, GJD2, IGKC, IGKV1-5, GPER, ADORA2A, CYTSA, CRISP1, UBE2J1, ACRV1, PIM2, CYP2E1, CD38, MGAT2, TRAPPC9, TXNDC5, FER1L4, IGHD, SPAG4, HES2, NLE1, PLA2G2A, IGKV4-1, PRDM1, ORAI2, GPR144, IGH@, SAA1, GAST, IGHG1, SAA2, MANF, TRAM2, IGLV3-19, CDC42EP2, IGLJ3, HLA-DOA, C16orf3, DCAKD, ZC3H13, ST6GAL1, C12orf32, RYBP, C21orf91, SERPINB3, SERPINB, SLAMF7, IGK@, LOC100129250, ZNF215, KCNJ4, NR1I2, ATP2A3, CD300A, IL5RA, HSPA13, |

| | |
|---|---|
| | LZTS1, LDLR, ELF5, TTLL4, FCRL2, TRGV3, LOC10029005, TP63, WBP4, KCNJ14, C22orf24, GJA3, RABAC1, SCT, CSH2, DIP2C, TTC30A, C8orf30A, ZFYVE9, CHST11, CSH1, SLC22A2, CRYBB3, TARP, RAP2A, LOC91316, ICAM2, ICAM3, ARTN, GZMB, MBD2, MMP14, ZBTB25, HNRPDL, SLC26A4, MAST1, LAX1, CCND2, PLXDC1, NCK1, NPPC, C1QL1, C19orf6, TMCO3, FKBP11, ZER1, MED1, FAM69A, FFAR2, LRRC36, SEC14L4, IGHA1, ITM2C, SEC14L1, FAM46C, ANKRD36, ZFP36L1, XBP1, BCL2, ZNF473, MEG3, HSF2BP, RFPL3, NCRNA00081, ZBP1, ABCA12, PDK1, CAPN6, PAOX, SPANXA1, SPANXA2, TNFRSF6B, NOS1, LRRC41, CEP135, TRIM26, GJB4, CD180, SOD2, ERP44, IGHV3-23, LOC1002, CD19, ADCY9, PPBPL2, RTEL1, C19orf57, AOX1, ABCC3, LIME1, ATP6V0A1, CMA1, SSR4, TAPBPL, SSR3, IGLV2-18, MFSD2B, F2RL3, THRA, SLC6A2, PGLYRP1, KIAA0746, MYL10, FGF12, APOBEC3F, MRPS31, CRADD, KRT81, GATA2, APOA2, DMXL2, CSNK2A1, NANS, SLC2A5, GPR44, IGHA2, CFB, SRRM1, DNAJC1, MYST4, FKBP1B, P11, NMNAT2, POU2AF1, ZNF440, DLL3, TNFRSF17, C2, BASP1, RRP9, EGFL8, NEBL, PPT2, OGFRL1, RPAIN, GABRR1, TRIM31, TFAP2A, NEU2, KIR2DL2, ASCL3, MAVS, PPY, DERL1, SLC38A7, DRD4, ITGB4, TCL1A, CPLX3, LMAN1L, UMOD, TCL1B, SRF, IRAK4, IRAK3, GCKR, TCP10L, ZNF695, CNR1, PCYT1B, SSX2, D4S234E, HERPUD1, PTPN2, EFEMP2, TRGC2, LOC652494, IGJ, SLC6A13, SPPL2B, MED13L, IDH3A, ATF5, PHF2, DNAJB9, ZNF419, TXNDC15, SPCS3, CD79A, EAF2, CACNA1F, LDOC1, GPR31, CACNA1A, SEL1L, NXPH3, C1orf38, NPEPL1, CLTA, SEC24A, NXPH4, CYTIP, C20orf19, OGDHL, HR44, PRDX4, MAGOH2, RHOQ, EPS15L1, ARHGAP17, PAWR, AMN, SKAP1, FRMD1, SLC1A4, CTTN, ALAS2, PRMT2, GRWD1, PAK3, TPP1, PROZ, DLG4, C2orf34, DAZ1, DAZ2, DA, ITIH3, DENND5B, SLC43A1, SLC1A1, TNIP3, HPD, RHOH, VCPIP1, CSHL1, SOX10, SOX14, CHST2, NCR1, NCR3, EML3, IFNAR2, KIR3DL2, LOC72778, FAM82B, BTG2, STXBP6, SIGLEC5, COMMD3, SPATS2L, VPREB1, WBP11, TPSG1, MSX2, SNN, COL9A3, N4BP1, OXCT1, KLK6, SLC12A3, L, NF1, DTX3, DPYSL4, ISOC2, RHAG, CYBA, PNOC, LOC10013386, KCNN3, C1RL, CENPT, IRF4, SCAND2, ADAMDEC1, IFNA17, SH3BP2, TCP11L1 |
| Sun_a (46) | RPL27A, NACA, RPL34, RPS3, LOC51035, IFRD2, PAK1, PEX7, SLC2A1, INHA, SEC31L1, STX1A, MST1R, FUT3, CXCL3, GPC3, IRX5, CORO1A, MAP1A, TRIO, MS4A1, P2RY6, IGLL1, GAP43, SPOCK2, PDE7A, PTPN9, PTPRCAP, RNASE2, BTK, CXCL12, KIAA0274, MAP3K12, IRF2, ALDH1A1, POU5F1, ARHGDIB, GTF2H2, CNN3, AMFR, PRKACB, DBP, FUCA1, ALDH9A1, RFTN1, DAXX |
| Sun_b (43) | ENDOD1, SLC7A1, CARHSP1, PGLS, NOL3, PYGL, MEP1B, CHKA, HIG2, BACE2, CUBN, FLJ21511, IL8, POU2AF1, CCND2, CASK, HLF, ZFP64, ATP8A2, PLCL2, DTNB, SPAST, MANEA, PELI2, ADRBK2, ITM2A, Sept11, POLR2J2, ZNF551, NGDN, MARK1, OGFRL1, IL16, CDC2L5, SSBP2, ABCC4, BACH2, LGALS8, SDCCAG3, CASR, EDG2, DTX3, RUNX3 |
| Tang (19) | DOCK9, RRM2, AURKA, HOPX, PRC1, GPR116, NKX2-1, TTC37, CDKN3, COL4A3, IFT57, ATP8A1, C1orf116, CYP2B6 , CYP2B7P1, DPP4, HSD17B6, MBIP, SLC35A5 |
| Tomida1_a (23) | WEE1, LYPLA1, PTN, ARG1, MST1, ACTR3, MYC, SSBP1, SNRPB, ARCN1, SERPINE1, PTP4A3, TITF1, SFTPC, NAP1L1, SERPINB1, FOSL1, THBD, CTNND1, NICE-4, CCT3, SPRR1B, COPB |
| Tomida1_b (17) | ESTs, KRT5, PTP4A3, SPRR1B, LOC339324, MYST4, SPARCL1, IGJ, EIF4A2, ESTs, ID2, THBD, MGC15476, ZFP, COPB, ZYG, CACNA1I |
| Tomida1_c (12) | NICE-4, WEE1, SSBP1, WFDC2, ACTA2, G22P1, MST1, PHB, DRPLA, SNRPB, GJA, SFTPC |
| Tomida2 (46) | API5, ARMC8, BRWD1, C11orf73, C6orf62, CACYBP, CKAP5, COIL, COL9A1, CXorf56, E2F6, EIF2S1, EIF2S2, ELF5, FIP1L1, GOLGA7, HNMT, HNRPH3, HSD17B12, IFT52, LIME1, MTERFD1, OR11A1, PDCL3, PGRMC1, PICALM, PRPF4B, PSMD10, PSMD12, RANBP9, RBM34, RBMX, RNF170, RPP40, SEC22A, SMARCE1, SRP9, STK38, TADA2L, TIPRL, TMEM14B, UBE2V2, UMPS, VBP1, VDAC3, ZBTB33 |
| Wilkerson (208) | AOF2, MYL6B, TMSL8, PODXL2, HSF2, TTLL4, MARCKSL1, MDK, CHKA, TRIM28, STOM, CASP1, GM2A, SQRDL, C1S, SERPINB1, THBD, PRKCH, MALL, CSTA, ADAM23, FOXE1, CHST7, SOX2, TUFT1, TP63, SLC9A3R1, ALDH3A1, GLI2, ALDH3A2, PHC2, PTP4A2, CD63, CRIP2, SLC43A3, TNS3, CD47, |

| | |
|---|---|
| | C1orf123, TRIM8, FAM46A, SLC16A4, ARHGDIB, DPP4, BIRC3, TNFRSF14, ALOX5, FMNL1, HLA-DRA, TPCN1, RASGRP3, BATF, PTPRE, ITGAM, HLA-DPA1, ASAH1, NRP1, CSF2RA, SLC39A8, ARL6IP5, IQSEC1, NKX2-1, MDFIC, RAB20, SERPINA1, TCIRG1, IL15RA, CASP4, RARRES3, TK2, ST3GAL5, IL21R, TMED5, MICAL1, ACSL5, TMC5, SH3BP5, SMPDL3A, RBPJ, IL32, SP100, SGPP1, ANXA5, GZMB, ARHGEF3, GZMH, A2M, ARRB2, PKP1, PERP, DSC3, SLC6A8, VSNL1, JUP, ABCC5, TRIM29, SERPINB5, GPX2, PIR, SIAH2, NSDHL, DLG1, DSG3, NCBP2, KLF5, RAB40B, MARK1, SFRS10, PRKCI, CELSR2, ADH7, TMEM14A, DRG1, KRT17, TFRC, NDUFB5, GLTP, EIF2B5, RNF7, PITX1, HMGCS1, CLCA2, ST6GALNAC2, LCMT2, FANCE, TMPRSS4, HRAS, BDH1, RAE1, TPD52L1, SQLE, PTPRZ1, SREBF2, ALG3, S100A9, CXCL1, SERPINB4, EPHA2, S100A8, SERPINB3, TAX1BP3, S100A2, DHRS3, IL8, CABC1, NUP210, FAM125B, CDK5RAP2, LPIN1, SLC29A1, ODC1, USP13, TACSTD1, KIAA1598, EFHD1, FNBP1L, ZNF239, PLEKHB1, KIAA0182, PHF17, BICD2, CBR1, TXN, SLC12A6, ZNF74, ACTL6A, WASF1, TSN, ASCC3L1, WDR12, NIPSNAP1, PDCD2, ILF2, LAPTM5, MNDA, DPYD, ALOX5AP, SLCO2B1, NCF4, CYBB, GLRX, MS4A6A, C11orf49, ABI2, SPAG5, RBBP7, GJB5, GJB3, SFN, GPR87, AHNAK2, AKR1C3, PRKX, HNMT, GMFG, ME1, TALDO1, IL4R, PDZK1IP1, ALDH1A3, DSE, MMP10, VDR, CAPZB, FNBP1, ENPP4, SH2B3, DOCK10, SDC1 |
| Wistuba (31) | TK1, CEP55, PBK, RAD54L, NUSAP1, RRM2, KIAA0101, ORC6L, RAD51, CENPM, SKA1, CENPF, KIF11, PTTG1, CDC2, DTL, PLK1, CDCA3, ASF1B, TOP2A, ASPM, CDCA8, MCM10, FOXM1, CDC20, CDKN3, BIRC5, DLGAP5, KIF20A, BUB1B, PRC1 |
| Xie (59) | H3F3A, HNRNPK, HSPD1, DEK, CBX3, NBPF15, ATP1B1, NDUFAB1, NUP153, CTSH, KIAA0101, PLOD2, LSM5, NBN, MTIF2, PSMA4, TBCA, CLPX, HNRNPA2B1, DBT, SNRPG, SNRPA1, DDX17, MRPL3, HMGB2, CYCS, NFIB, ATP5I, RGL1, CBLB, ZMYM2, TUBA1B, BLVRA, LARP1, MED13L, IDS, GNS, ATP6V0A1, FCF1, ABCC10, SFTPB, BTF3, CYB5A, YPEL5, UBE2J1, HIGD1A, NUSAP1, CCDC90B, CCDC59, OLA1, FBXO38, DENR, ANGEL2, N4BP2L2, MCM4, EZH1, KIAA0240, SIDT2, DNAJC21 |
| Zhu (15) | ATP1B1, TRIM14, FAM64A, FOSL2, HEXIM1, MB, L1CAM, UMPS, EDN3, STMN2, MYT1L, IKBKAP, MLANA, MDM2, ZNF236 |
